# Cost-effectiveness of respiratory syncytial virus vaccination for older adults: a modelling analysis

**DOI:** 10.64898/2026.07.08.26357577

**Authors:** Victoria L. Oliver, Julian B. Carlin, Yingying Wang, Violeta Spirkoska, Adrian Marcato, Kylie S. Carville, Robert Moss, David J. Price, Patricia T. Campbell, Jodie McVernon, Natalie Carvalho

## Abstract

**Background:** Evidence of the effectiveness and cost-effectiveness of new vaccines that reduce the burden of respiratory syncytial virus (RSV) in older populations is emerging. The reported cost-effectiveness of these vaccination strategies varies substantially across different settings. This study assessed the cost-effectiveness of older adult-targeted RSV vaccination strategies in the Australian context and compared findings with published evaluations.

**Methods:** We developed an individual-based dynamic transmission model of RSV infection, linked to a clinical pathways and cost-effectiveness model. We modelled different adult vaccination strategies for the general population and the Indigenous population, and present incremental cost-effectiveness ratios (ICERs) as cost per quality-adjusted life year gained, from a healthcare system perspective. Deterministic and probabilistic sensitivity analyses explored drivers of cost-effectiveness and sensitivity of findings to uncertainty in parameter estimates.

**Results:** Vaccinating the general population of older adults in Australia was not found to be cost-effective at a dose price of 100 AUD, but was found to be cost-saving for Indigenous adults, given the higher disease burden in this population. Individual drivers of ICERs in our setting were dose price, hospitalisation incidence and mortality, however conclusions about cost-effectiveness were robust to joint parameter uncertainty.

**Conclusions:** The cost-effectiveness of vaccinating adults against RSV depends on many uncertain and context-specific quantities. Strategies that target high risk populations were found to be cost-effective in Australia due to the larger avertable burden.

**Key Summary Points:** *Why Carry Out This Study?:* Respiratory syncytial virus (RSV) is a significant cause of respiratory illness and death, particularly in older adults, individuals with medical conditions, and Indigenous populations. The reported cost-effectiveness of RSV vaccination strategies varies substantially across different settings. With the Australian Government beginning to fund RSV vaccines from May 2026, this study aimed to provide timely, Australia-specific evidence on which vaccination strategies offer the greatest benefit relative to their cost.

*What Was Learned from the Study?:* A structured literature review confirmed substantial variability in how costs and benefits of RSV vaccination are evaluated across settings, underscoring the importance of context-specific evidence and transparent reporting when informing national vaccination policy decisions. In our analysis, vaccinating Indigenous Australians aged 60 years and over was found to save money for the health system while also preventing hospitalisations and deaths, primarily because of the high burden of RSV in this population. Vaccinating the broader older adult population was not found to be good value for money at the vaccine prices and willingness-to-pay thresholds assumed in this study. The most influential drivers of cost-effectiveness were vaccine price and in-hospital case fatality rate - two factors that remain uncertain in the Australian context and warrant ongoing monitoring.

## 1. Introduction

Respiratory syncytial virus (RSV) is a significant cause of respiratory illness worldwide, ranging from mild upper respiratory tract infections to severe lower respiratory tract disease, with the greatest burden in children under five years of age [1]. RSV also causes substantial morbidity and mortality in older adults and in those with comorbidities, in whom lower respiratory tract infections can be especially severe [2]. Although RSV cases are actively monitored in several high-income countries including Canada [3] and the United States of America (USA) [4], legal notification is only mandated in a few, such as Australia since 2021 [5,6], and the United Kingdom (UK) since 2025 [7]. The differences in RSV surveillance across settings and over time contributes to variability in estimates of global incidence rates [8]. The adjusted pooled hospitalisation incidence rate from six studies in high-income settings (USA, Finland, and New Zealand) has been estimated at 347 (95% Confidence Interval (CI), 203–595) per 100,000 per year for adults aged ≥65 years, with the in-hospital case fatality rate (hCFR) estimated at 6.1% (95% CI, 3.3–11.0%) (8). Similar estimates were reported in a meta-analysis of RSV burden among adults aged ≥60 years in Europe, where hospitalisation incidence ranged from 193 (95% CI, 125–304) per 100,000 per year in the Netherlands to 414 (95% CI, 322–514) in Denmark, with hCFR ranging from 6.73% (95% CI, 4.63–9.69%) in Spain to 10.14% (95% CI, 4.91–19.79%) in Switzerland [9]. Data for low- and middle-income countries, particularly adult populations, are scarce, with additional challenges in estimating disease burden, such as variable healthcare access and limited diagnostic capacity, beyond those faced by high-income countries [10,11].

A higher clinical burden and more serious outcomes are observed in individuals with comorbidities such as chronic respiratory diseases, including asthma and chronic obstructive pulmonary disease, who face greater likelihood of hospitalisation, longer hospital stays and higher case fatality rates [2,12–14]. Other conditions associated with increased risk of severe RSV disease include cardiac disease, immunocompromising conditions (e.g. malignancy, HIV or solid organ transplant), chronic metabolic disorders (e.g. diabetes), chronic neurological disorders, chronic liver or kidney disease and obesity [15,16], with RSV likely to exacerbate these underlying conditions [17]. Globally, Indigenous peoples including Aboriginal and Torres Strait Islander Australians are also at an increased risk of severe RSV disease, and more likely to be hospitalised and experience longer hospital stays [18–21].

Several vaccines against RSV disease have become commercially available including Arexvy (GlaxoSmithKline) and Abrysvo (Pfizer) protein subunit vaccines, and an mRNA vaccine, mResvia (Moderna). The cost-effectiveness of current protein subunit RSV vaccines for adults has been evaluated in several settings, with substantial variability in findings. Research from Japan suggests that vaccination of adults ≥60 years is cost-effective compared to no vaccination using a willingness to pay (WTP) threshold equivalent to ∼30,000 United States dollars (USD) per quality-adjusted life-year (QALY) [22,23]. In contrast, an analysis in the USA found that vaccination of ≥60 or ≥65 year olds was not cost-effective using a WTP threshold of 100,000 USD/QALY [24].

In 2024, Abrysvo and Arexvy were approved by the Therapeutic Goods Administration in Australia, and recommended by the Australian Technical Advisory Group on Immunisation (ATAGI) for adults aged ≥75 years, adults aged ≥60 years with medical risk conditions, and Indigenous adults aged ≥60 years [15]. From May 2026, the Australian Government is funding Arexvy for all adults aged ≥75 years, and Aboriginal and Torres Strait Islander adults aged ≥60 years, through the National Immunisation Program (NIP)[15,25]. The Pharmaceutical Benefits Advisory Committee (PBAC) has also provided a positive recommendation for Arexvy (with a price reduction) for adults aged 60 to 74 years [26]. Recent studies have evaluated the cost-effectiveness of RSV vaccination for adults aged ≥75 years in Australia and found that Arexvy would need to be priced at $150 Australian dollars (AUD) or less to be considered cost-effective in all socio-economic strata at a willingness to pay threshold of $50,000 AUD per QALY gained [27,28]. However, there is limited evidence to understand the cost-effectiveness of RSV vaccination for Australian adults with elevated risk of severe outcomes, such as those with comorbidities or Aboriginal and Torres Strait Islander people. We extended a published model of RSV transmission and cost-effectiveness in infants [29] to evaluate the potential impact and cost-effectiveness of currently recommended [15] and alternative RSV vaccination strategies for older adults in Australia compared to no vaccination. This analysis was performed independently of national cost-effectiveness analyses conducted by PBAC.

## 2. Methods

### 2.1 Study design

We conducted a literature review to summarise the evidence available at the time of the modelling on the cost-effectiveness of RSV vaccination for older adults, followed by a cost-utility analysis to evaluate the cost-effectiveness of different vaccination programs for this population in the Australian setting. The cost-effectiveness analysis used a pipeline of three models we previously developed to assess the cost-effectiveness of infant RSV immunisation products [29]. These models included 1) a transmission model of RSV spread and vaccination roll-out, 2) a clinical pathways model that probabilistically simulated clinical endpoints given an infected individual’s age, risk status, and vaccination status, and 3) a cost-effectiveness model that calculated incremental costs and QALYs for each vaccination strategy compared to the current no-vaccination scenario. We evaluated four vaccination strategies compared with no-vaccination. For the general Australian population, we explored three main vaccination strategies: i) vaccination of adults ≥75 years of age only; ii) vaccination of adults ≥75 years of age and at-risk adults ≥60 years of age; and iii) vaccination of adults ≥60 years of age. For the Indigenous population we explored one vaccination scenario, iv) vaccination of adults ≥60 years of age. In a scenario analysis, we explored intermediate strategies between ii) and iii) by reducing the age-eligibility threshold (for adults not-at-risk) to 70, or 65 years.

### 2.2 Literature review

The search strategy was designed based on the PICO framework as follows: P (population): non-pregnant adults aged ≥18 years; I (intervention): RSV vaccination with any product; C (comparator): any comparator; O (outcome): incremental cost-effectiveness ratio (ICER) or net monetary benefit (NMB). MEDLINE (Ovid) was searched on 14^th^ July 2025 and all studies published since database inception were screened. The details of search terms used along with inclusion and exclusion criteria are provided in Table 1 of Additional file 1. Articles were screened based on title and abstract to identify potentially relevant studies. The full texts of all studies remaining after this first screening stage were retrieved and screened to identify studies meeting inclusion criteria. Reference lists of included articles were reviewed to identify other potentially relevant articles. These articles were screened based on their abstract and then full text as described above. A data extraction template was used to summarise model design elements, key inputs, and cost-effectiveness findings from each article. A qualitative synthesis of the extracted data was performed to summarise major commonalities and differences across studies in terms of methods and findings.

### 2.3 Modelling transmission

The details of the transmission model are included in Carlin et. al. [29]. In short, the transmission model was a dynamic, individual-based model of 100,000 individuals with demographic characteristics representative of the general Australian population (hereafter ‘general population model’), or the Indigenous Australian population (hereafter ‘Indigenous population model’). Demographic data was sourced primarily from the Australian Bureau of Statistics (ABS; see Additional File 1 for further detail).

We assumed that RSV spreads via age-stratified population-level mixing of individuals, with contact matrix as estimated by Mistry et al. [30]. The model captured an individual’s progressive acquisition of natural immunity following repeated exposures to RSV via a tiered susceptible-exposed-infectious-recovered-susceptible (SEIRS) compartmental structure [31,32].

The transmission and clinical pathways models were calibrated simultaneously such that the modelled incidence of RSV-related healthcare presentations in the no-vaccination scenario conformed with historical Australian data from the 2018–2019 RSV seasons. We chose these years due to the impact of COVID-19 on RSV circulation, and more recent data were not available. Specifically, we used national hospitalisation data with an RSV-coded diagnosis, whether this was the primary admission reason or otherwise. These data (including intensive care unit (ICU) admissions, and deaths) were provided by the Department of Health, Disability and Ageing using Admitted Patient Care data supplied by the Australian Institute of Health and Welfare (AIHW). The data were provided as aggregated numbers, with suppression applied, in five-yearly age groups, by month for hospitalisations, and by year for ICU admissions and deaths.

### 2.4 Modelling clinical and health-related quality of life outcomes

In Australia, RSV became a notifiable disease in 2021. As such, the number of recorded hospitalised cases in 2018 and 2019 is likely an under-estimate of the hospitalisation burden. RSV detection has been shown to be approximately 1.5 times higher when combining serology with PCR testing [33]. As RSV serology is not commonly used for diagnosis in Australia, we multiplied the raw counts of RSV-coded hospitalisations by a factor of two to account for both reduced sensitivity of PCR testing alone and incomplete testing of patients with respiratory illness [33]. We did not apply this under-ascertainment factor to the number of recorded ICU admissions or deaths, as testing is more routinely performed for individuals at these more severe clinical endpoints.

As shown in Figure 1, the clinical pathways model probabilistically estimated the clinical endpoint of each RSV infection, given each individual’s age and risk-status. Explicitly, we modelled seven mutually exclusive endpoints: no medical care is sought, a general practitioner visit (GP), emergency department consultation with no admission to a hospital ward (ED), admission to a hospital ward without further progression, admission to an ICU, death following admission to an ICU, and death following admission to a hospital ward without admission to an ICU. From our Australian data we computed the age-dependent transition probabilities of progression from admission to a hospital ward to more severe clinical outcomes of ICU and/or death. The output of the clinical pathways model is stochastic; due to the variable chains of infections in the transmission model, and that the outcome of each infection is probabilistically simulated. For all strategies, we simulate outcomes from the clinical pathways model 10,000 times (500 different clinical outcomes from 20 different transmission simulations). Mathematical details and a full discussion of the data sources and assumptions for the clinical pathways model are in Additional file 1.

**Figure 1:**
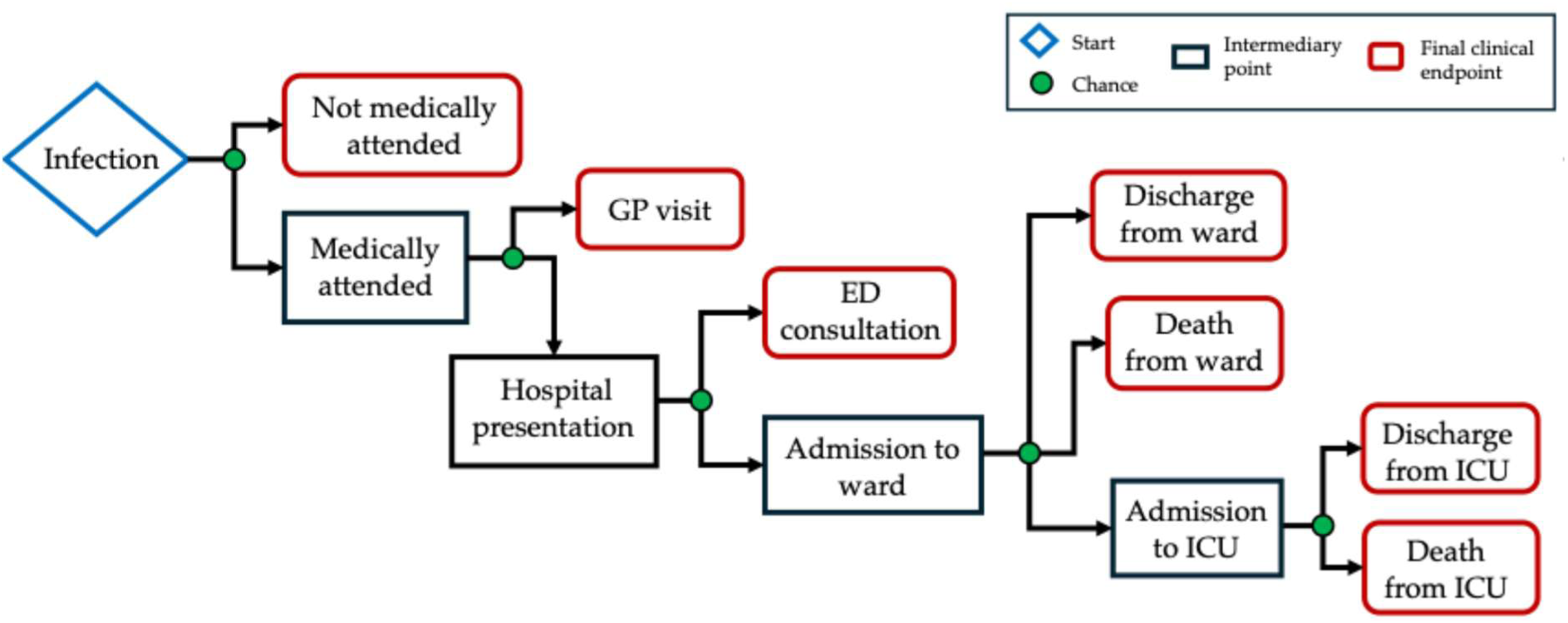
Clinical pathways model structure.

Adults with risk conditions, including cardiac disease, chronic respiratory, neurological and metabolic disorders, chronic kidney disease and obesity are at increased risk of severe outcomes from RSV [15]. Although the ABS National Health Survey (2023) reports the number of individuals with long-term conditions, it does not enable estimates for each condition that account for multi-morbidity [34]. Estimates for the prevalence of having at least one risk condition are presented in Table 2 of Additional file 1. In short, we assumed that roughly 20% of adults 60 years of age have at least one risk condition, rising steadily with age to roughly 40% of adults 75 years of age, agreed to in consultation with ATAGI.

Australian data on the relative risk (RR) of hospitalisation for adults with at least one risk condition (hereafter “at-risk adults”) aged ≥60 years were not available. We derived a RR estimate from a prospective community study in the USA over 2004–2016, that presented RRs of hospitalisation for adults with chronic obstructive pulmonary disease, congestive heart failure, asthma, diabetes and immunocompromising disorders individually [14]. We estimated a weighted average RR of hospitalisation, adjusting for the prevalence of each condition, using the raw data reported in Belongia et al. [14]. The estimated RR of hospitalisation for the at-risk cohort compared to the not-at-risk cohort was 2.29. We assumed, in the absence of other evidence, that this RR applies across all medically-attended clinical endpoints for adults in the general population model, regardless of age.

For the Indigenous population we treated all adults aged ≥60 years of age as at-risk for the purposes of vaccine eligibility and disease outcomes. RSV-coded hospitalisations, using AIHW-supplied data from 2012–2019, show an incidence rate ratio (IRR) of 2.9 (95%CI 1.7–5.1) and 1.5 (95%CI 0.8–2.6) for Indigenous Australian adults aged 60–64 and ≥65 years, respectively, compared to the non-Indigenous population [35]. An independent analysis of Queensland data from 2018–2023 shows an IRR of 3.1 (95%CI: 2.7–3.6) in the Indigenous compared to the non-Indigenous population, aggregated across all ages [36]. As a pragmatic middle-ground, we determined that a single IRR of 2.9 is appropriate for Indigenous adults aged ≥60 years. To capture this in the clinical pathways model, we back-calculated a multiplier to the probability of each medically attended clinical endpoint, such that we observed an IRR of 2.9 in our no-vaccination scenarios. Further detail is provided in Additional file 1.

Age-specific utility values were based on Australian population norms to reflect background age-specific QALY (in the absence of RSV infection) and were applied to both the general population and Indigenous population models [37]. Health related quality of life decrements for non-admitted infections (those with GP visits or ED presentations) and hospitalisations (general ward and ICU) were based on EQ-5D-5L derived utility weights as reported in a study in Canada [38]. We assumed that the duration of illness for non-admitted cases was five days, while the duration for hospitalised cases was based on length of stay data from National Hospital Cost Data Collection dataset (NHCDC; round 26, 2021–2022). For deaths, we calculated QALY decrements as the total remaining discounted quality-adjusted life expectancy at age of death using general population life tables for Australia. Life expectancy tables for the general population were applied to both the general population and Indigenous population models.

### 2.5 Modelling vaccination impacts

The clinical trial vaccine efficacy (VE) endpoints of RSV vaccines for older populations do not map directly to the modelled clinical endpoints [39–42]. As such, we did not attempt any head-to-head comparisons of the relative effectiveness of different products. In consultation with ATAGI, we determined one set of product-agnostic VE inputs to evaluate in the model, informed by evidence from relevant clinical trials and emerging real-world effectiveness studies for each of the products to date [43,44]. The initial efficacy for each clinical endpoint is listed in Table 1 for not-at-risk and at-risk adults (including Indigenous adults). These base case VE values were varied by ±20% as part of the sensitivity analyses. We did not ascribe any protection against infection (whether asymptomatic or symptomatic), as there is no direct evidence for this mode of action in RSV vaccines to date. As a result, we observed the same infections in both the no-vaccination and vaccination strategies, allowing direct computation of the individual-level averted clinical burden due to vaccination. Data indicate declining VE over two to three seasons, and in the absence of longer-term data, we assumed that the initial VE against each endpoint waned linearly to zero from the date of vaccination over three years [45]. We simulated vaccine delivery to be uniformly distributed seasonally, between February and July in the first year of the two-year time horizon, with a year-round delivery strategy explored as an alternative scenario analysis. For all strategies, we assumed a total uptake of 60% would be reached in the eligible population whether a year-round or seasonal program was in place (i.e. same number of doses delivered in both strategies), consistent with historic influenza vaccination uptake in these age groups [46]. As others have done [38,47–49] we did not model adverse events due to vaccination as data from clinical trials suggest these are minor and transient [40,41].

**Table 1:** Input parameters for modelling health outcomes and costs.

| Parameter | Base case value (lower estimate, upper estimate; distribution used for PSA) | Source |
| --- | --- | --- |
| **Clinical endpoint probabilities** |  |  |
| GP visit given infection | Age- and risk-status-dependent (0.9× base case, 2× base case; triangular) | Base: see Additional file 1; Lower/upper: assumption |
| ED presentation given infection | Age- and risk-status-dependent (0.9× base case, 2× base case; triangular) | Base: see Additional file 1; Lower/upper: assumption |
| Hospitalisation given infection | Age- and risk-status-dependent (0.9× base case, 2× base case; triangular)<sup>a</sup> | Base: see Additional file 1; Lower/upper: assumption |
| ICU admission given hospitalisation | Age- and risk-status-dependent (0.9× base case, 2× base case; triangular) | Base: see Additional file 1; Lower/upper: assumption |
| Death given ICU admission or hospitalisation | Age- and risk-status-dependent (0.9× base case, 5× base case; triangular) | Base: see Additional file 1; Lower/upper: assumption |
| **Disutility weights** |  |  |
| Outpatient cases (GP or ED visit) | 0.16 (± 50%; gamma) | [38] |
| General ward hospitalised cases | 0.41 (± 50%; gamma) | [38] |
| ICU hospitalised cases | 0.60 (± 50%; gamma) | [38] |
| **Duration of disutility** |  |  |
| Outpatient cases | 5 (3–8; beta) days | Assumption |
| General ward hospitalised cases | 4.4 (1.6–6.2; beta) days | NHCDC and [50] |
| ICU hospitalised cases | 9.8 (7.4–13.6; beta) days | NHCDC and [50] |
| **Initial vaccine efficacy against GP visit** |  |  |
| Not-at-risk adults | 70% (±20%; normal) | Product agnostic estimate based on clinical trial data and ATAGI advice |
| At-risk adults | 65% (±20%; normal) | Product agnostic estimate based on clinical trial data and ATAGI advice |
| **Initial vaccine efficacy against ED presentation, hospitalisation, ICU admission, and death** |  |  |
| Not-at-risk adults | 80% (±20%; normal) | Product agnostic estimate based on clinical trial data and ATAGI advice |
| At-risk adults | 75% (±20%; normal) | Product agnostic estimate based on clinical trial data and ATAGI advice |
| **Vaccine program costs** |  |  |
| Vaccine dose costs, AUD | 100 (80, 250; uniform)<sup>b</sup> | Base: CDC pricing [51] scaled down to 43.6%; Lower: base case – 20%; Upper: CDC pricing [51] |
| Administration costs, AUD | 36.35 (31.05, 42.85; gamma) | Base: mean of lower and upper; Lower: MBS item 82205; Upper: MBS item 23 |
| Wastage | 5% (0-10%; beta) | Assumed based on national guidelines [52]. |
| **Outpatient healthcare costs** |  |  |
| Cost of each GP visit for RSV, AUD | 41.40 (±50%; gamma) | MBS item 23 |
| Number of GP visits per case of RSV | 2 (1-3; discrete) | Assumption |
| Cost of a virologic test per outpatient case of RSV, AUD | 28.65 (±50%; gamma) | MBS item 69494 |
| Cost of over-the-counter drug per outpatient case of RSV, AUD | 16.15 (±50%; gamma) | PBS item 3192B |
| Cost per non-admitted ED presentation, AUD | 724 (±50%; gamma) | NHCDC round 26 (2021–2022) (national average cost of non-admitted ED) |
| **Hospitalisation healthcare costs** |  |  |
| Per admitted ED presentation, AUD | 1,335 (±50%; gamma) | NHCDC round 26 (2021–2022) (national average cost of admitted ED) |
| Per non-ICU hospitalisation, AUD | 7,980 (±50%; gamma) | See note<sup>d</sup> |
| Per ICU hospitalisation, AUD | 25,490 (±50%; gamma) | See note<sup>e</sup> |

ATAGI: Australian Technical Advisory Group on Immunisation; CDC: Centers for Disease Control and Prevention; ED: Emergency Department; GP: General Practice; ICU: Intensive Care Unit; MBS: Medicare Benefits Schedule; NHCDC: National Hospital Cost Data Collection; PSA: Probabilistic Sensitivity Analysis; QALY: Quality-Adjusted Life Year.
<sup>a</sup> Lower estimate used for two-way sensitivity analysis was 0.5× base case to represent an assumption of no ascertainment bias in national surveillance data.
<sup>b</sup> For the two-way sensitivity analysis, dose price was varied between 0 and 400 AUD
<sup>c</sup> Scale factor based on reported average differences in medicines wholesale prices between the United States and Australia [53]
<sup>d</sup> NHCDC data using the following AR-DRG: E62A (Respiratory Infections and Inflammations, Major Complexity), E62B (Respiratory Infections and Inflammations, Minor Complexity), E70A (Whooping Cough and Acute Bronchiolitis, Major Complexity) and E70B (Whooping Cough and Acute Bronchiolitis, Minor Complexity). Final cost represents an average across these DRGs, weighted by the number of episodes reported in the NHCDC per DRG.
<sup>e</sup> NHCDC data using the following AR-DRG: E41A (Respiratory System Disorders W Non-Invasive Ventilation, Major Complexity) and E41B (Respiratory System Disorders W Non- Invasive Ventilation, Minor Complexity). Final cost represents an average across these DRGs, weighted by the number of episodes reported in the NHCDC per DRG.

### 2.6 Measurement of costs

We adopted a formal healthcare system perspective and included vaccination program costs and healthcare costs for medically attended cases of RSV infection. As the dose price for RSV vaccines in Australia is unknown, we used a hypothetical dose price of 100 Australian dollars (AUD) based on U.S. Centers for Disease Control and Prevention (CDC) prices moderated down by an estimate of the average price differential of medicines in Australia compared to the USA [51,53]. Inputs and sources for modelling wastage and vaccine administration costs are shown in Table 1. Programmatic costs related to vaccine introduction, supply and storage, or surveillance were not included. GP visit costs included the cost of two consultations, one pack of over-the-counter medication and one RSV test. The costs of ED presentations and hospitalisations were from NHCDC (round 26, 2021–2022). The selection of diagnosis related groups (DRGs) was informed by a study of the annual cost burden of children with RSV in Australia [50] and prior PBAC submissions [54]. Costs are reported as 2025 AUD.

### 2.7 Cost-effectiveness and sensitivity analyses

ICERs were calculated, representing the additional costs per QALY gained for each vaccination strategy compared to no vaccination over a two-year horizon, with QALY losses associated with deaths projected over a life-time horizon. A 5% discount rate was applied to future costs and benefits in line with national PBAC guidelines [52]. Explicit cost-effectiveness thresholds are not available for Australia. Studies have suggested that 50,000\ AUD/QALY gained may be indicative of a positive recommendation from the PBAC [55,56]. Others have suggested 15,000 AUD/QALY gained may be more reflective of a threshold used for decision-making on vaccines and other preventative medicines in Australia [57,58]. We compared ICERs to each of these thresholds when interpreting the cost-effectiveness of vaccination strategies, acknowledging that decision-makers may consider higher or lower thresholds alongside other considerations, such as equity, budget impact and implementation feasibility.

To explore the sensitivity of results to uncertainty in input parameters, we conducted one-way and two-way deterministic sensitivity analyses. These analyses were conducted for vaccination strategies aligned to recommendations from ATAGI and involved varying one or two parameters to the upper or lower estimate of a range of plausible values while keeping all other parameters at base case values. A probabilistic sensitivity analysis (PSA) was conducted in which the 10,000 model simulations sampled multiple parameters from a range of plausible values according to a probability distribution (See Table 1 for a list of parameters varied in the sensitivity analyses). Given the uncertainty in vaccine dose price, this parameter was kept fixed at the base case value (100 AUD) in the PSA, with an alternative analysis conducted with the dose price sampled from within the uncertainty range according to a uniform distribution. The probability of the vaccination strategy being cost-effective compared to no vaccination, across a range of potential WTP thresholds, was estimated by calculating the proportion of simulations which were equal to or below a given threshold between 0 to 150,000 AUD/QALY (varied in 100 AUD increments).

## 3. Results

### 3.1 Findings from literature review

The database search yielded 1,452 unique articles. After title and abstract screening, 66 articles were identified for full text screening. Of these, 17 were included. A further article was identified from bibliographic screening to yield a total of 18 included articles (see Figure 1 of Additional file 2 for a flow chart of articles included at each stage of screening). Across the 18 articles we identified 35 unique analyses, defined as being based on a unique model, study setting or vaccine product. All analyses evaluated cost-effectiveness of RSV vaccination for older adults in high-income settings, including several countries in Europe [31,47,48,59–61], North America [24,38,62–68], and Asia [22,23,55]. Figure 2 gives a graphical representation of articles and analyses identified, where analyses have been grouped according to comparator (no vaccination or next best alternative RSV vaccination strategy), type of economic metric reported (ICER or vaccine price required for positive NMB) and assessment (cost-effective or not compared to locally-relevant WTP thresholds).

**Figure 2:**
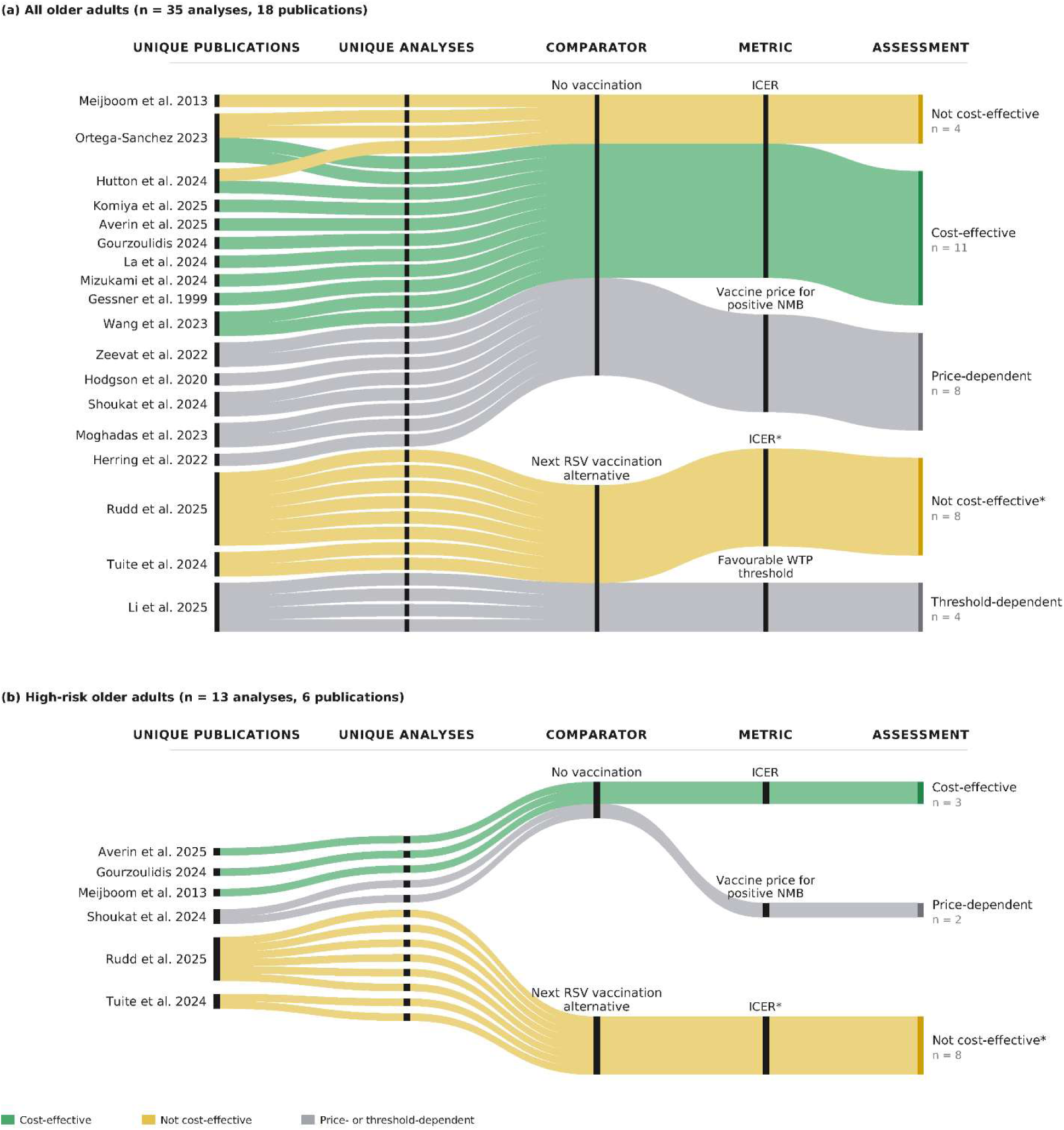
Sankey charts illustrating breakdown of findings from published studies evaluating the cost-effectiveness of vaccination of adults aged ≥60 or ≥65 years (top) or adults aged ≥60 or ≥65 years with elevated risk. “Unique Analyses” were defined as being based on a unique model, study setting, vaccine brand. “Comparator” strata represent whether the vaccination strategy was compared to no vaccination or the next best RSV vaccination strategy in adults. “Metric” strata represent the format for presentation of results: i) an incremental cost-effectiveness ratio (ICER), ii) the vaccine price at which positive net monetary benefit is achieved, or iii) the willingness to pay threshold at which the vaccination strategy is preferred over alternatives. “Assessment” strata represent whether the vaccination strategy was deemed cost-effective. ICER: incremental cost-effectiveness ratio: NMB: net monetary benefit; WTP: willingness to pay * ICER and cost-effectiveness assessment are based on comparison to next best alternative RSV vaccination strategy

Vaccination of adults aged ≥60 or ≥65 years (regardless of risk status) was evaluated in all 35 analyses, with 23 analyses comparing this strategy to no vaccination, and 12 conducting a sequential cost-effectiveness analysis in which this strategy was compared to the next best alternative RSV vaccination strategy for older adults. Among the 23 analyses adopting no vaccination as the comparator, eight modelled the maximum justifiable price to achieve a positive NMB. Estimates ranged from 17 USD for an analysis in the Netherlands with a WTP threshold equivalent to 22,000 USD/QALY [60] to approximately 300 USD for an analysis in the USA with a WTP threshold of 106,000 USD/QALY [67]. Of the remaining 15 analyses, 11 found vaccination of adults aged ≥60 or ≥65 years to be cost effective compared to no vaccination at their base case dose price (ranging from 50–295 USD; median = 200 USD) using locally relevant WTP thresholds [22–24,47,59,62,64,68], with the remaining four analyses finding this strategy not to be cost-effective (at base case dose prices).

Thirteen analyses evaluated a strategy of vaccinating adults aged ≥60 years with elevated risk of severe disease, five of which compared this strategy to no vaccination. Three of these five analyses found this strategy to be cost-effective at base case dose prices [47,59,61], with the remaining two finding positive NMB at dose prices up to 119–127 USD (assuming a WTP threshold equivalent to 36,000 USD/QALY) [38].

Commonly reported drivers of cost-effectiveness included vaccine price, hospitalisation incidence, mortality risk or incidence, and vaccine efficacy and waning. There was significant variability in the values used for these parameters across published analyses, with no discernible relationship between any one parameter and overall cost-effectiveness conclusions. See Table 2 of Additional file 2 for a summary of key input parameters and findings of published analyses presented alongside our study for comparison.

### 3.2 Model calibration

The transmission and clinical pathways models were calibrated to match the main features of RSV disease in Australia. Figure 3 shows that the model recovered the monthly hospitalisations per 100,000 population in five age groups, once the observed data were multiplied by an under-ascertainment factor of two. The numbers of deaths in the model were also consistent with 2018–2019 Australian data: we simulated on average 460 deaths per year in adults ≥60 years of age, when our results are scaled up to the number of Australians ≥60 years of age, compared to the 404 and 474 deaths attributed to RSV in our national dataset in 2018 and 2019 respectively. Our model estimated the annual incidence of infection (regardless of clinical endpoint) per 100,000 adults ≥60 years of age is 37,720 (5^th^ to 95^th^ percentile 36,301 to 39,447). Details of model parameters fit as part of the calibration procedure are included in Additional file 1.

**Figure 3:**
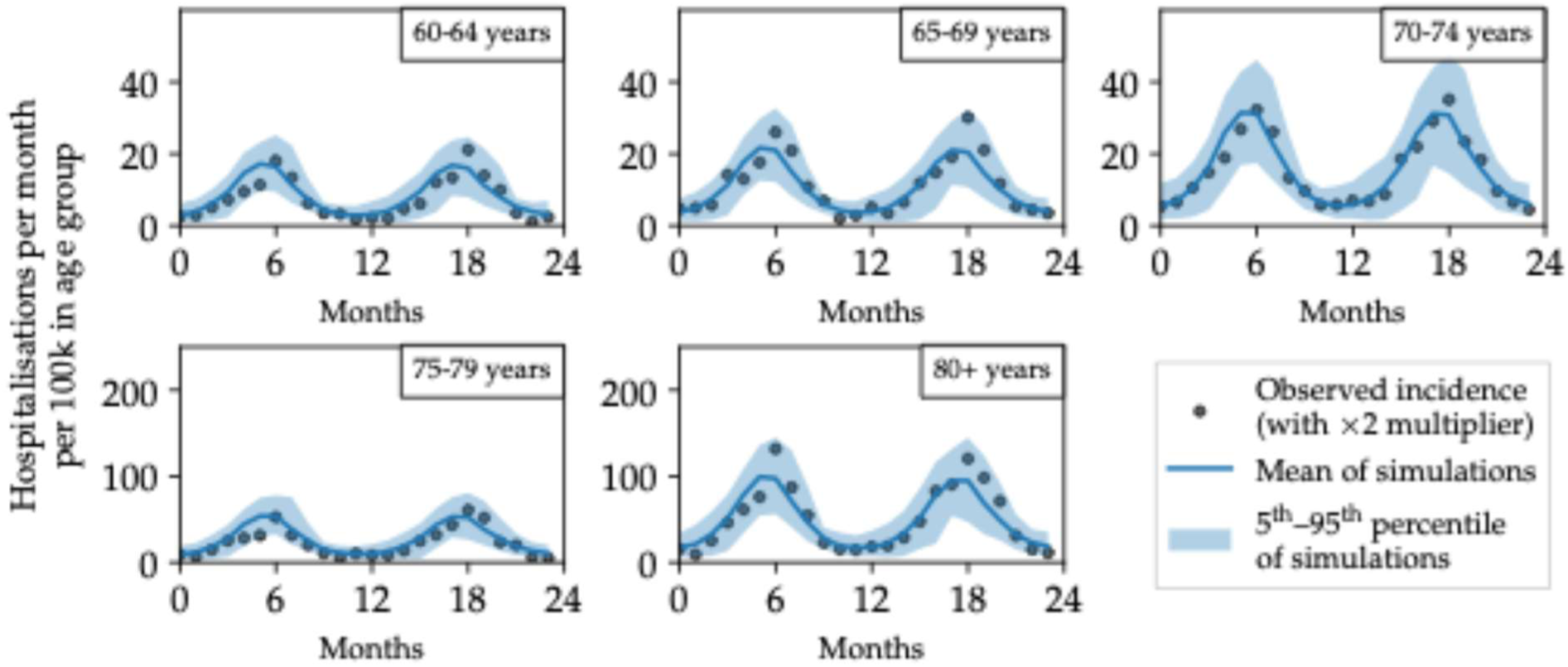
Monthly hospitalisations per 100,000 in each age group from the transmission model and the age-specific infection-hospitalisation rate (blue curve is the mean, blue band is the 5^th^ to 95^th^ percentile from 100 transmission model simulations), compared to observed monthly hospitalisations in 2018–2019 (black points, with a multiplier of two applied).

### 3.3 Base case impact and cost-effectiveness

The impact of vaccination on hospitalisations and deaths is shown in Figure 4, with impact on the number of GP visits, ED presentations, and ICU admissions provided in Additional file 2 (Table 2). In the general population model, we found that without vaccination an estimated 469.7 (5^th^–95^th^ percentile: 391–551) hospitalisations and 8.3 (1–20) deaths were expected per year in a population of 100,000 adults aged ≥60 years. Given an overall vaccination uptake of 60%, the strategy with the largest impact on health outcomes was when adults aged ≥60 years were vaccinated seasonally (Feb through July), with 135.6 (94–184) hospitalisations and 2.5 (0–10) deaths averted each year (29% reduction for both). With this strategy, the numbers needed to vaccinate (NNV) to avert one hospitalisation and one death were 483 and 26,220 respectively (Table 3 of Additional file 2). In the Indigenous population model, we found that without vaccination, an estimated 1363.4 (1151–1585) hospitalisations and 24.3 (2–56) deaths were expected per year in a population of 100,000 adults aged ≥60 years. Given 60% uptake of vaccination among Indigenous adults ≥60 years, 362.0 (248–483) hospitalisations and 6.5 (1–26) deaths were averted each year (27% reduction). This equated to a NNV of 104 and 5,815 to avert one hospitalisation and one death, respectively (Table 3 of Additional file 2).

**Figure 4:**
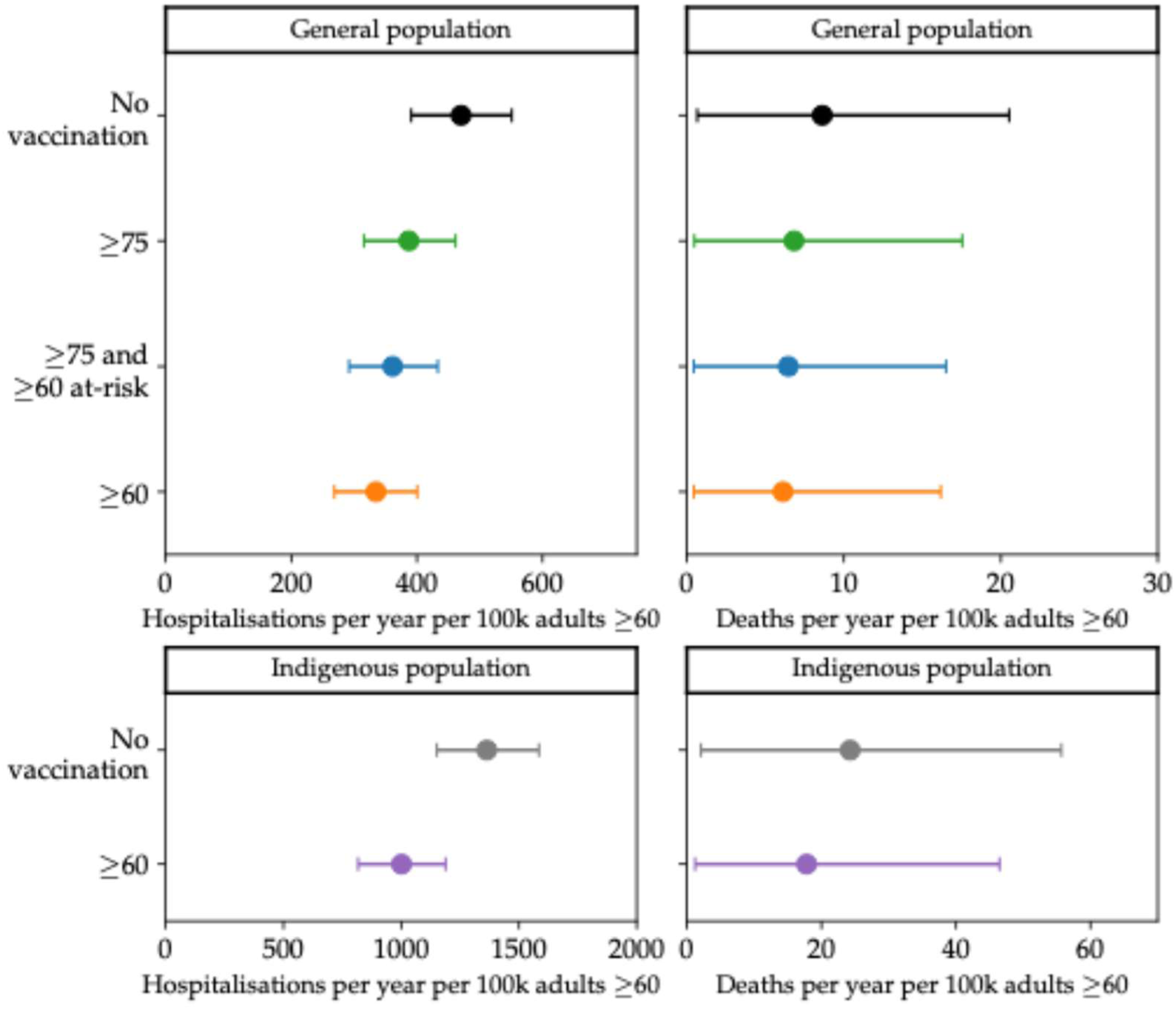
Impact of vaccination on hospitalisations and deaths averaged over two years for the general population (top) and Indigenous population (bottom) models. All vaccination scenarios shown have doses delivered seasonally (Feb through July) in the first year of the simulation, with 60% uptake in the age-eligible population.

**Table 2:**
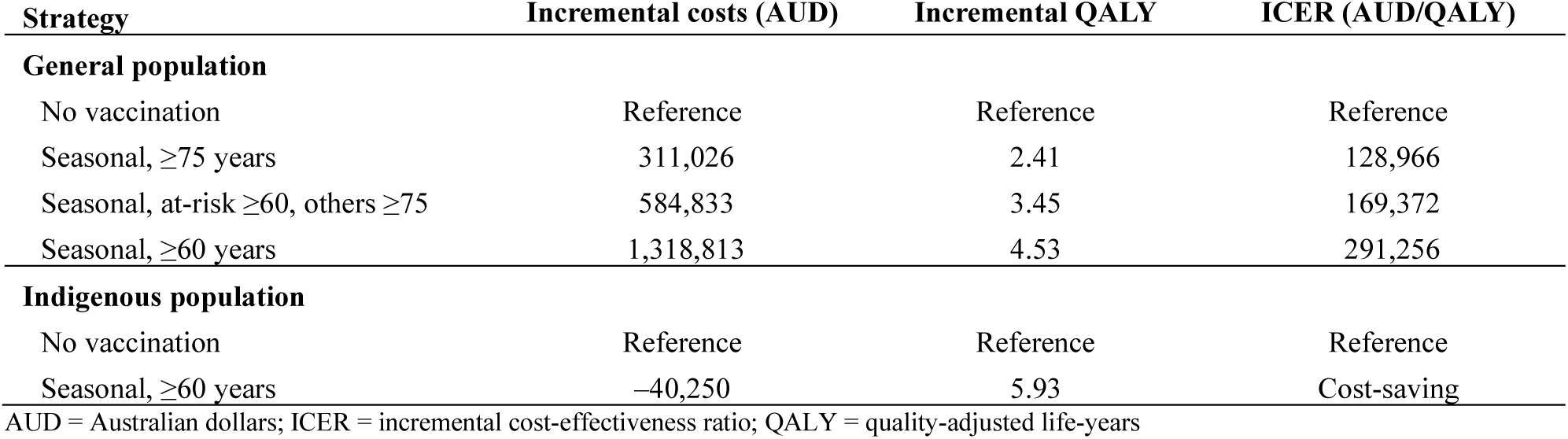
Base case cost effectiveness results. AUD = Australian dollars; ICER = incremental cost-effectiveness ratio; QALY = quality-adjusted life-years

| Strategy | Incremental costs (AUD) | Incremental QALY | ICER (AUD/QALY) |
| --- | --- | --- | --- |
| **General population** |  |  |  |
| No vaccination | Reference | Reference | Reference |
| Seasonal, $\geq 75$ years | 311,026 | 2.41 | 128,966 |
| Seasonal, at-risk $\geq 60$, others $\geq 75$ | 584,833 | 3.45 | 169,372 |
| Seasonal, $\geq 60$ years | 1,318,813 | 4.53 | 291,256 |
| **Indigenous population** |  |  |  |
| No vaccination | Reference | Reference | Reference |
| Seasonal, $\geq 60$ years | -40,250 | 5.93 | Cost-saving |

Results from analysis of alternative vaccination strategies are provided in Table 2 of Additional file 2. In the general population model, lowering the age-eligibility cut-off from 75 years (down to 70, 65 or 60 years) averted more disease, at the cost of more total doses being delivered. Delivering vaccines as part of a year-round strategy averted fewer hospitalisations on average than an equivalent seasonal strategy, despite the same number of doses being delivered to achieve a 60% uptake rate overall, see Table 2 of Additional file 2.

Results from the base case cost-effectiveness analysis are shown in Figure 5 and Table 2. Cost-effectiveness planes show the incremental costs and QALYs associated with each simulation of the transmission and clinical pathways model. Simulations without deaths averted by vaccination can be seen as the dense cloud of points closest to the y-axis, which account for approximately a half to two-thirds of the 10,000 simulations. The clouds of data points extending to the right represent simulations where at least one death is averted in the vaccination strategy compared to no vaccination. This morphology of points is an artefact of our simulation having 100,000 agents; the density of the ‘no deaths’ cloud and spread of the ‘tail’ would be reduced in an Australian-sized population model. This artefact does not affect the headline cost-effectiveness analysis results, which are based on the clinical burden averaged across all simulations, which is well calibrated to the clinical burden observed in the Australian population. Based on the mean outcomes of all simulations, vaccination of age-eligible adults in Australia resulted in an incremental discounted cost to the health system ranging from 311,026 AUD (for vaccination of adults aged ≥75 years) to 1,318,813 AUD per 100,000 population (vaccination of adults aged ≥60 years) over two years. The QALY gains were predominantly (∼79%) driven by averted deaths and ranged from 2.41 (vaccination of adults aged ≥75 years) to 4.53 (vaccination of adults aged ≥60 years) per 100,000 population. ICERs for these vaccination strategies ranged from 128,966 AUD/QALY to 291,256 AUD/QALY.

**Figure 5:**
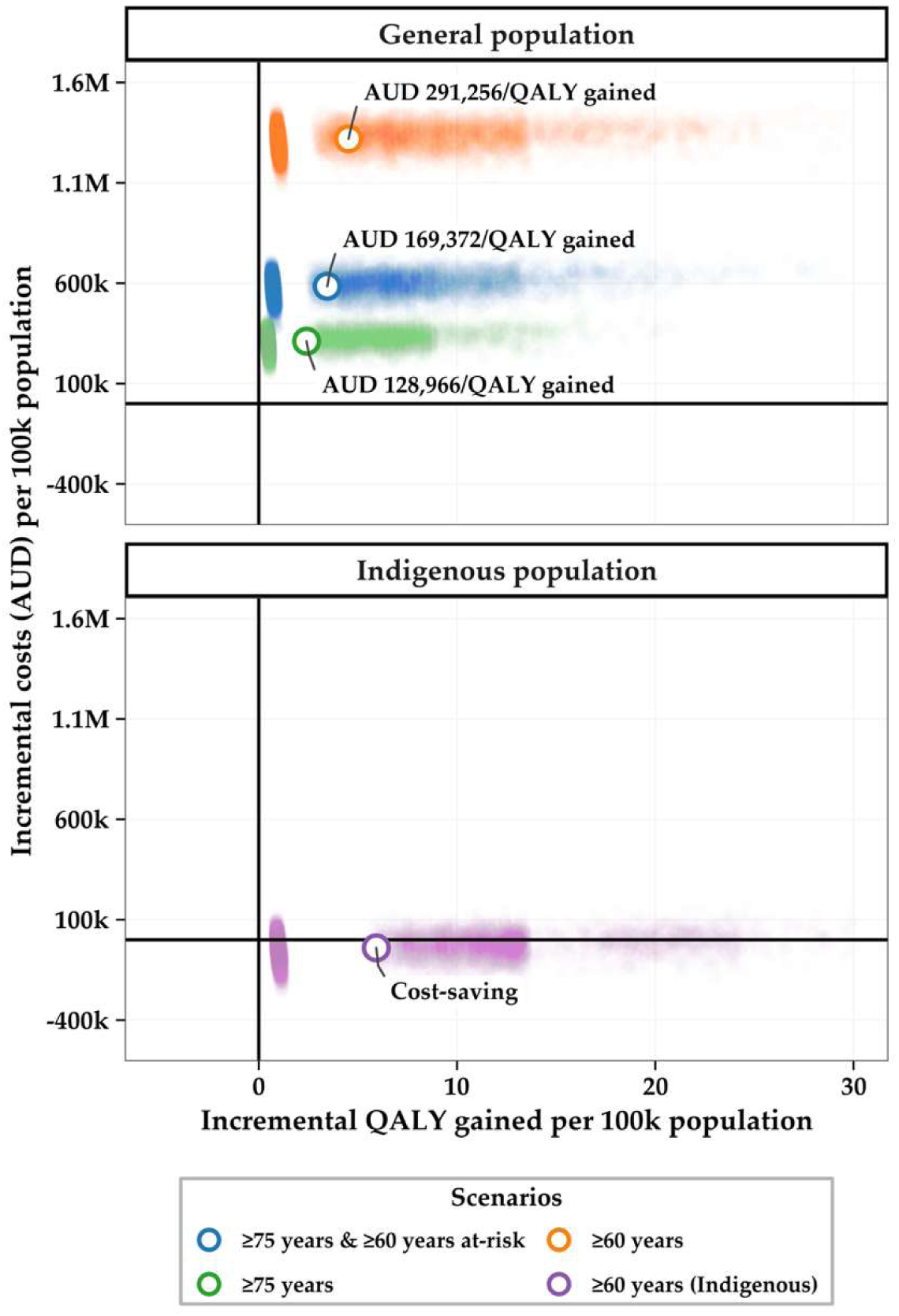
Incremental costs and QALYs modelled from 60% uptake seasonal vaccination strategies for the general population (top) and the Indigenous population (bottom) compared to no vaccination. Coloured dots show data for each simulation output from the clinical pathways model (spread of data represents stochastic variation in infection dynamics and the clinical pathways model). The visible gap between the cluster closest to the y-axis and the clouds extending to the right indicate the difference between no deaths averted and at least one death averted in the simulated population. Open dots show the mean incremental cost and QALY across all outputs, with estimated incremental cost-effectiveness ratio. AUD: Australian dollars; QALY: quality-adjusted life-year

Vaccination of Indigenous adults aged ≥60 years was estimated to yield 5.93 QALY per 100,000 population and result in a net discounted saving to the health system of 40,250 AUD over two years.

### 3.4 Uncertainty analysis

A one-way deterministic sensitivity analysis was conducted for two strategies reflecting ATAGI recommendations [15]: i) vaccination of adults aged ≥75 years along with adults with medical risk conditions aged ≥60 years and ii) vaccination of Indigenous adults aged ≥60 years (Figure 2 of Additional file 2). Vaccine dose price had the largest effect on ICERs for both strategies. General population vaccination strategies were not cost-effective across the range of parameter values explored, except the hCFR. At the highest estimate for hCFR (equivalent to roughly 8.5% of hospitalised cases in adults aged ≥60 years), the ICER fell just below a threshold of 50,000 AUD/QALY. At the highest dose price explored (250 AUD), the ICER for vaccination of Indigenous adults was 74,327 AUD/QALY gained. This vaccination strategy was either cost-saving or had an ICER of less than 12,500 AUD/QALY for all other parameter values explored.

A two-way deterministic sensitivity analysis explored the impact of the two most influential parameters on vaccination strategies for the general population (vaccine dose price and the hCFR) or Indigenous population (vaccine dose price and hospitalisation incidence) (Figure 6). At the base case dose price of 100 AUD, vaccination of adults aged ≥75 years along with medically at-risk adults aged ≥60 years was cost-effective (against a threshold of 50,000 AUD/QALY) when the hCFR was more than four times greater than our assumed base case value. A dose price of less than 50 AUD would be necessary for this vaccination strategy to be cost-effective at our base case assumption for hCFR. At our base case assumption for hospitalisation incidence, vaccination of Indigenous adults aged ≥60 years was cost-saving or cost-effective at dose prices of up to 223–290 AUD (based on thresholds of 15,000 and 50,000 AUD/QALY, respectively). If we assumed no under-ascertainment bias in reporting of RSV hospitalisation, dose prices of less than 205 AUD would be required for this vaccination strategy to be cost-effective at a threshold of 50,000 AUD/QALY.

**Figure 6:**
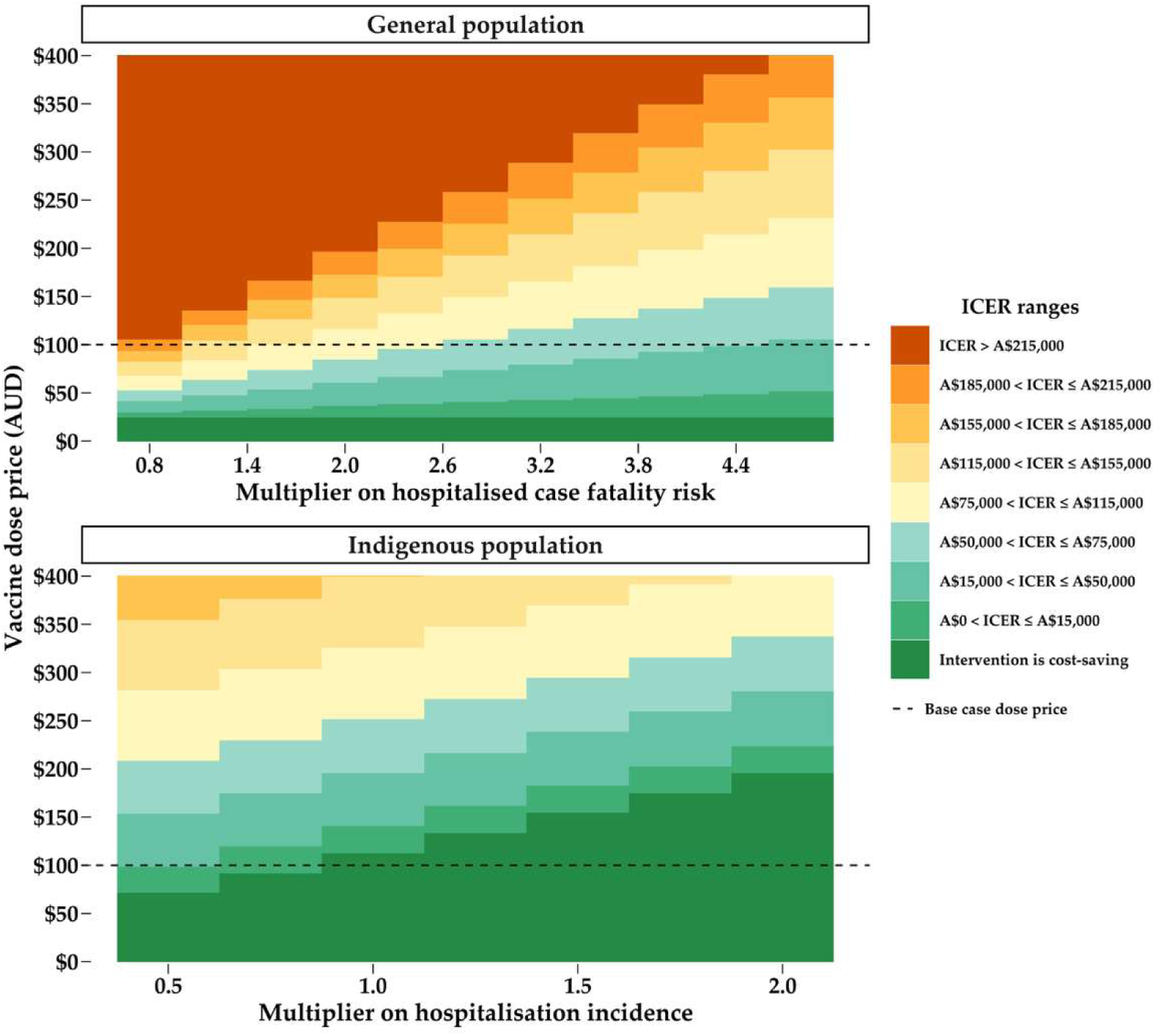
Results from the two-way sensitivity analysis of vaccination strategies for the general population (top) and the Indigenous population (bottom) compared to no vaccination. For the general population, vaccination of adults aged ≥75 years and at-risk adults aged ≥60 years was modelled. For the Indigenous population, vaccination of adults aged ≥60 years was modelled. Units of x-axes represent multipliers applied to base case values of hCFR (top) and hospitalisation incidence (bottom). AUD: Australian dollars; ICER = incremental cost-effectiveness ratio; QALY: quality-adjusted life-year

Cost-effectiveness acceptability curves from the probabilistic sensitivity analysis are shown in Figure 7, with the cost-effectiveness planes shown in Figure 2 of Additional file 2. At a dose price of 100 AUD, the general population vaccination strategy had a probability of being cost-effective of 0.05 and 0.26 for WTP thresholds of 15,000 and 50,000 AUD/QALY, respectively. This vaccination strategy had <0.4 probability of being cost-effective up to the highest WTP threshold explored (150,000 AUD/QALY), regardless of whether dose price was fixed (at 100 AUD) or sampled from a distribution (uniformly between 80 and 250 AUD).

**Figure 7:**
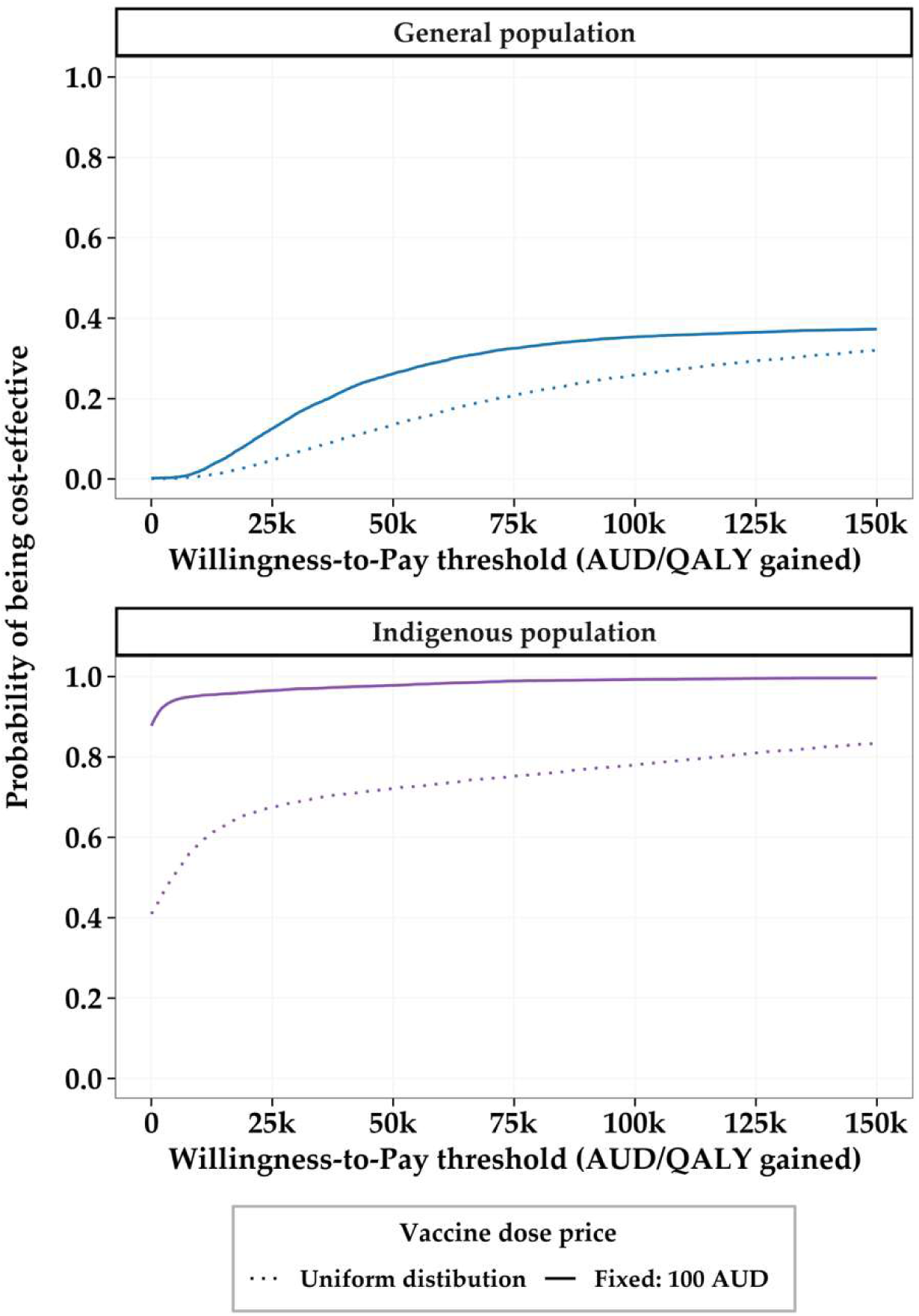
Cost-effectiveness acceptability curves from probabilistic sensitivity analysis of vaccination strategies for the general population (top) and the Indigenous population (bottom) compared to no vaccination. For the general population, vaccination of adults aged ≥75 years and at-risk adults aged ≥60 years was modelled. For the Indigenous population, vaccination of adults aged ≥60 years was modelled.AUD: Australian dollars; QALY: quality-adjusted life-year

At a fixed dose price of 100 AUD, vaccination of Indigenous adults (aged ≥60 years) had a probability of being cost-effective of 0.96 and 0.98 for WTP thresholds of 15,000 AUD and 50,000 AUD, respectively. The probability of cost-effectiveness was lower (<0.8 across all WTP thresholds) if uncertainty in dose price was included in the PSA.

## 4. Discussion

To our knowledge, this is the first study to report on the cost-effectiveness of RSV vaccination for Indigenous adults and those with medical risk-based indications for vaccination in Australia. We found vaccination of Indigenous adults aged ≥60 years would likely be cost-saving in the base case. Cost-effectiveness was robust to uncertainty in almost all parameter inputs except vaccine dose price, which remains highly uncertain in Australia. Our base case dose price estimate was derived from US CDC vaccine price lists [51], scaled to 46% to reflect estimates of the differences in drug prices between Australia and the USA [53]. When CDC prices were used without scaling, the ICER for vaccination of Indigenous adults exceeded a commonly used WTP threshold of 50,000 AUD/QALY. Our literature review did not identify other studies evaluating the cost-effectiveness of selective RSV vaccination for Indigenous adults in any setting. A few studies evaluated vaccination for other adult populations with elevated risk, such as persons with medical risk conditions [47,59,61], or residents of long-term care facilities [38]. As in our analysis, these studies generally found risk-based vaccination strategies to be cost-effective compared to no vaccination, depending on dose price.

Our findings suggest that at the base case vaccine price considered, strategies involving vaccination of age-eligible adults in the general population (regardless of risk) are unlikely to be cost-effective based on a commonly used threshold of 50,000 AUD/QALY. This finding was consistent across a range of vaccination strategies, including those with different age-eligibility cut-offs (≥60 years or ≥75 years) and when younger adults (≥60 years) with medical risk conditions were eligible for vaccination alongside adults age ≥75 years. Our findings were also consistent across a plausible range of uncertain input parameters. These findings differ from recent studies from Australia, which found RSV vaccination to be cost-effective for adults aged ≥75 years at the same threshold of 50,000 AUD/QALY [27,28]. A major difference between our model and these published studies relates to VE parameters. As described in Section 2.5 we assumed that VE against death started at 80% (75% for at-risk individuals) and waned linearly to zero over three years post-vaccination. This assumption results in minimal protection during the third RSV season post-vaccination, and as such we set our time horizon to two years to capture the bulk of the averted burden due to vaccination. On the other hand, Paul and colleagues [27] assumed a VE against hospitalisation and ICU admission (and therefore also death) of 94.1% in the first year, 64.2% in the second year, and 43.3% in the third year, which would drive more favourable cost-effectiveness results. There are limited real-world studies of RSV VE over multiple years, which results in different model assumptions. As more evidence arises this uncertainty will decrease.

Approximately three quarters of the studies from our literature review which compared vaccinating adults aged ≥60 or 65 years to no vaccination found that vaccination was cost-effective using locally-relevant WTP thresholds. Compared to these analyses, our model uses some parameter estimates which would be more favourable for cost-effectiveness, such as a relatively low dose price (68 USD compared to a median of 200 USD in published analyses) and high incidence of hospitalisation (491 per 100,000 persons ≥60 years compared to a range of 46 – 413 in published analyses). However, other model parameters were less favourable, such as the hCFR used in our clinical pathways model, which was at least 2.8-fold lower than those used in studies identified in our review.

The hCFR used in our model was derived from the total number of hospitalisations and deaths from hospitals that are coded as RSV-related (via ICD-10-AM codes for principal or additional diagnoses) in Australia from 2018 and 2019. As described earlier, we applied a multiplier of two to the raw number of hospitalisations recorded, due to an estimate of under-ascertainment in Australian hospitals [33]. We did not apply this multiplier to the raw number of RSV-related deaths, as the ascertainment bias is likely much lower at that endpoint. Most of the studies in other settings applied an ascertainment bias multiplier (typically between 1.3 and 2) to both hospitalisation numbers and deaths. Additionally, most of the other studies only consider hospitalisations as RSV-related if that was the principal coding, perhaps leading to increased under-ascertainment of hospitalisations compared to deaths, inflating their estimates of hCFR compared to ours. We acknowledge that there may be some degree of uncertainty associated with estimates of the RSV-mortality rate in Australia, highlighting the importance of strengthening RSV surveillance systems. However, while the magnitude of the ICERs was sensitive to the hCFR, the ultimate assessment of whether RSV vaccination for a general population of older adults would be considered cost-effective in Australia was largely unchanged across of range of plausible hCFR values.

Compared to the difference in hCFR, the overall mortality rate used in our model (which encompasses hospitalisation incidence and hCFR) was more similar, but still lower than that used in most other studies. Our estimates of the NNV to avert one death were 2.8 – 11.5 times greater than published studies which reported these results. Given that a significant proportion of QALY gains was driven by averted deaths, this suggests that the relatively low incidence of RSV-related mortality in Australia may limit the cost-effectiveness of vaccination strategies in which all older adults (regardless of risk status) are eligible for vaccination.

Our literature review highlighted that a wide range of parameter estimates have been used across publish cost-effectiveness analyses of RSV vaccination. This illustrates the uncertainty and variability in the data and assumptions underlying modelled economic evaluations, underscoring the importance of context-specific models and transparency in reporting of methods and assumptions. Further, reporting results which can facilitate comparisons between models from different settings should be encouraged. For example, estimates of NNV can enable comparisons across cost-effectiveness models on the basis of disease burden and vaccine effectiveness inputs regardless of setting-specific economic parameters (such as healthcare costs and willingness to pay). However, we identified only a few studies from our literature review which reported NNV.

Our sensitivity analyses showed that dose price, followed by the hCFR, were the most influential parameters on the ICER of Australia’s recommended RSV general population vaccination strategy (vaccination of adults ≥75 years along with adults aged ≥60 years with medical risk conditions). When considering a WTP threshold of 50,000 AUD/QALY, this strategy may be considered cost-effective if the risk of death among hospitalised cases was five times greater than our assumption based on national surveillance data. However, under these conditions the ICER would still exceed the more conservative threshold of 15,000 AUD/QALY, which has been proposed as reflective of decision-making on vaccines in Australia [54,57]. Explicit WTP thresholds are not available for Australia and, in addition to cost-effectiveness, decision-makers will consider vaccination strategies against multiple implicit and explicit criteria, such as feasibility of implementation, budget impact and equity considerations. Further, it should be noted that this analysis has been conducted independently from the health technology assessment process undertaken by PBAC, which underpinned the Australian Government’s decision to fund Arexvy for older adults under the NIP.

Our modelled outcomes have significant stochastic variability. This arises from two main factors: the model is an individual-based model of 100,000 individuals, of whom roughly 25% are over 60 years of age (for the general population model), matching Australia’s demographics. The number of modelled infections per year in this age group varies from 7,707 to 8,375 (5^th^ to 95^th^ percentile). The second factor is that the clinical pathways model simulates subsequent severity outcomes, which are relatively low probability events. Thus, the number of rare outcomes in adults aged ≥60 years, e.g. deaths, varies from 0 to 3 (5^th^ to 95^th^ percentile, per 100,000 individuals) in any individual simulated year. While the range of outcomes is broad, the mean result is well-estimated by averaging across all the simulations for each strategy.

We acknowledge limitations in our study. Firstly, while our structured literature review supports an understanding of how the methods, parameter inputs and findings of our analyses compare to those reported in the literature, a systematic search was not undertaken. Thus, the articles identified in this review do not represent an exhaustive list of relevant analyses. Secondly, our general population model generalises risk conditions to a singular “at-risk” flag, which likely underestimates risk for some individual conditions while overestimating it for others. If more granular data were available as to the RR of different conditions, and the prevalence of those conditions accounting for multimorbidity, we would be able to narrow the at-risk flag to those conditions which significantly increase the risk of severe RSV disease. This data availability limitation extends to other parameters in our model, such as hospitalisation incidence and hCFR, for which reliable estimates are lacking due to historical under ascertainment of RSV-related hospitalisations and changing testing accessibility and behaviours over time. Finally, death rates specific to the Indigenous population were used in the transmission model to ensure an accurate representation of the demographics for that subpopulation of Australia (for whom life expectancy is lower compared to non-

Indigenous Australians). However, when modelling QALYs gained from vaccination, general Australian population life tables were used to model life years lost in Indigenous adults. While this may overestimate QALY gains and the cost-effectiveness of RSV vaccination for this population, this approach is appropriate to ensure existing health inequities are not perpetuated by unequal valuation of outcomes in the Indigenous population compared to the general population. Equity considerations are addressed in this analysis through subgroup analysis; new methods such as distributional cost-effectiveness analysis may offer additional insights into the equity impact of RSV vaccination policies, but were beyond the scope of the current analysis.

This study found that vaccination of Indigenous Australian adults aged ≥ 60 years against RSV to be cost-saving or cost-effective, depending on dose price. However, in contrast to the findings of many published analyses, including recently published studies from Australia, vaccination of the general population of adults (regardless of risk status) was not found to be cost-effective in this analysis. Our work highlights the variability in methods and findings associated with economic evaluations of RSV vaccination, underscoring the importance of transparent and context-appropriate analysis and reporting.

## Declarations

### Availability of data and material

Data are provided within the manuscript and supplementary information file. Code and instructions on reproducing the results in the study are available at the following repository: https://gitlab.unimelb.edu.au/julian.carlin/older-population-rsv-modelling.

### Ethics approval and consent to participate

The model was parameterised using nationally aggregated patient data provided by the Department of Health, Disability and Ageing using Admitted Patient Care data supplied by the Australian Institute of Health and Welfare (AIHW). National Admitted Patient Care data (i.e. line by line unit records), collated by AIHW and supplied to the Department of Health, Disability and Ageing, were aggregated and suppressed by the Department before being provided to the authors. Approval for the use of these data for analyses and subsequent publication was granted by the Department.

### Consent for publication

Not applicable

### Authors’ contributions

Formal analysis (JBC, AJM, YW); Funding acquisition (JMcV); Investigation (JBC, AJM, YW); Methodology (JBC, AJM, YW, PTC, JMcV); Project administration (PTC, JMcV); Resources (JMcV); Software (JBC, YW, RM); Supervision (PTC, DJP, NC, JMcV); Validation-Verification (KSC, VLO, VS); Visualisation (JBC, YW); Writing – original draft (VLO, VS, JBC, AJM); Writing – review & editing (All). All authors have read and approved the final version.

## Data Availability

Data are provided within the manuscript and supplementary information file. Code and instructions on reproducing the results in the study are available at the following repository:
https://gitlab.unimelb.edu.au/julian.carlin/older-population-rsv-modelling.

https://gitlab.unimelb.edu.au/julian.carlin/older-population-rsv-modelling

## Acknowledgements

We thank the Australian Technical Advisory Group on Immunisation (ATAGI) secretariat and members, especially those within the RSV/respiratory working groups (chaired by A/Prof. Katherine Gibney) for provision of technical advice to support the development of this model. The ATAGI member list as of 2025 is available at https://www.health.gov.au/committees-and-groups/atagi/members.

We would like to particularly acknowledge Jocelynne McRae, Jean Li-Kim-Moy, Sanjay Jayasinghe and Bette Liu from the National Centre for Immunisation Research and Surveillance for working closely with us to provide access to AIHW-supplied data and in-confidence analyses.

