## Supplemental appendix 1 for "Cost-effectiveness of respiratory syncytial virus vaccination for older adults: a modelling analysis"

### Additional file 1: Further detail on methods

**Journal name:** Pharmacoeconomics.

Victoria L. Oliver[1]*, Julian B. Carlin[2]*, Yingying Wang[1]*, Violeta Spirkoska[2], Adrian Marcato[2], Kylie S. Carville[2,3], Robert Moss[1], David J. Price[1,2], Patricia T. Campbell[2], Jodie McVernon[2]^, Natalie Carvalho[1]^

*Joint first authors
^ Joint senior authors

[1] Melbourne School of Population and Global Health, University of Melbourne, VIC, Australia
[2] Department of Infectious Diseases, The University of Melbourne at the Peter Doherty Institute for Infection and Immunity, Victoria, Australia
[3] Victorian Infectious Diseases Reference Laboratory, The Royal Melbourne Hospital at the Peter Doherty Institute for Infection and Immunity, Victoria, Australia

**Table 1: Summary of literature search strategy**

| Research question framework | Search terms | Inclusion criteria (all criteria must be met for inclusion) | Exclusion criteria (any criteria can be met for exclusion) |
|---|---|---|---|
| **P (population):** Adults - other than pregnant - aged over 18 years women in any location; | No specific search terms used | Study population included adults who were eligible for vaccination based on age or medical risk status (other than pregnancy) | Studies on maternal and/or infant only vaccination strategies |
| **I (intervention)** RSV vaccination with any product | (Respirat* Syncyt* Vir* OR respiratory syncytial virus OR RSV) AND (vaccin* OR immuni* OR anti f-mab OR prefusion OR Abrysvo OR Arexvy OR mResvia OR mRNA) | Studies reporting on the impact of RSV vaccination using any commercially available or hypothetical vaccine | Studies on burden of disease only, no intervention considered |
| **C (comparator):** Any comparator | No specific search terms used | Studies which included any comparator to vaccination in the analysis | Studies without an identified comparator |
| **O (outcome):** A measure encompassing both the incremental costs and incremental benefits of vaccination compared to a relevant comparator | cost-effective* OR cost-benefit OR cost benefit OR cost effective* OR cost utility OR cost-utility OR cost consequence OR cost saving OR quality adjusted life year OR qaly OR disability adjusted life OR daly OR economic evaluation OR decision* OR mathematic* OR simulation* OR model OR Monte Carlo OR microsimul* OR simul* OR dynamic OR static OR agent based OR individual based | Studies reporting an incremental cost-effectiveness ratio or economically justifiable price | Studies reporting epidemiological outcomes only without economic outcomes<br><br>Vaccination program costs not factored into the analysis – e.g. only healthcare savings reported |

#### Demographic details and assumptions

The demographic sources and modelling choices for the general population strategies are described in the Appendix A of Carlin et al 2025[26]. For the Indigenous population strategies, we updated the age-sex distribution and probability of death at each age, according to Australian Bureau of Statistics data [69]. We show example age distributions for 100,000 individuals in the general population model and Indigenous population model in the left panel of Figure 1, while example age-at-death distributions are visualised in the right panel of Figure 1. We used the same contact matrix, see Appendix B.1 in Carlin et al 2025 [26], for both the general population and Indigenous population strategies.

**Figure 1: Age distribution and age-at-death distribution for the general population and Indigenous population. The age-at-death distribution only includes deaths that will happen in the next five years in the model.**

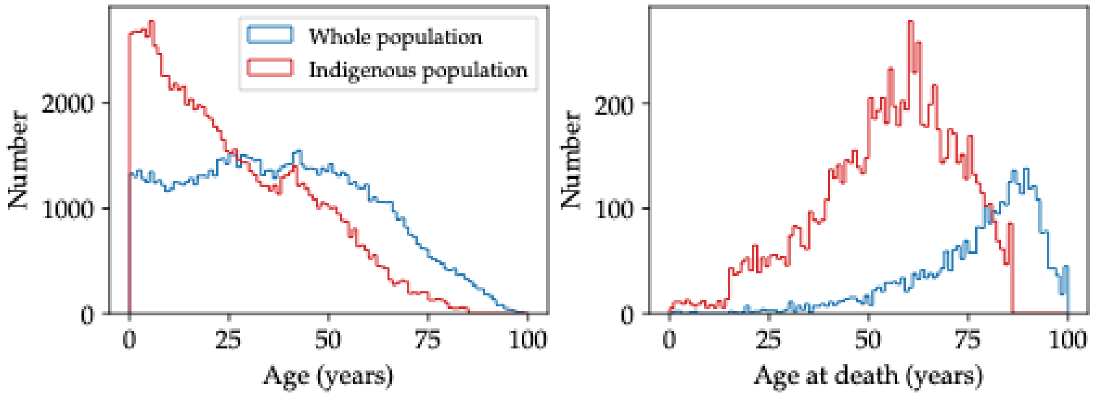


#### Calibration details

The calibration procedure and verification were described in Appendix C in the Supplementary Materials of Carlin et al 2025[26]. The one additional component necessary for the older population model was the infection to hospitalisation rate as a function of age for those over the age of 60. We found that a sigmoid with functional form $y = \frac{y_{max} + y_{min}}{2} + \frac{y_{max} - y_{min}}{2} \tanh[a\ (age\ +\ b)]$, with parameters 0.04, 0.001, 0.075, −86 for $y_{max}$, $y_{min}$, $a$, and $b$, respectively, adequately generated the appropriate hospitalisation incidence, given the infection incidence rate by age, see Figure 2 in the main text.

#### Clinical pathways details and assumptions

We used National Admitted Patient Care data (line unit records), collated by AIHW and supplied to the Department of Health, Disability and Ageing, which the Department aggregated and suppressed before being provided to the authors, to estimate appropriate age-specific transition probabilities between hospitalisation, ICU, and death.

The aggregated data contain, in five-yearly age groups up to 90 years of age, monthly for the years 2018 and 2019, the number of individuals who:

i) Are admitted to hospital, using International Statistical Classification of Disease and Related Health Problems, Tenth Revision, Australian Modification (ICD10AM) codes for principal diagnoses (J21.9) and principal or additional diagnoses (J12.1, J20.5, B97.4, J21.0);
ii) Are admitted to ICU, with ICD10AM diagnosis codes per above;
iii) Die, after being admitted to ICU, with ICD10AM diagnosis codes per above;
iv) Die, without admission to ICU, with ICD10AM diagnosis codes per above.

Using these numbers, we computed smoothed transition probabilities between hospitalisation, ICU and death endpoints as a function of age. The probability of death following an ICU admission did not appear to evolve with age, so we aggregated across all ages to find 113 deaths from 927 ICU admissions, giving a transition probability of 0.122 (95%CI: 0.102–0.145). The probability of ICU admission given hospitalisation was found to decrease linearly with age, with slope $-0.00107\ \text{years}^{-1}$, and ordinal offset 0.0384. The probability of death following hospitalisation (without ICU admission) increases with age according to an exponential with functional form $y = y_{min} + exp[-\lambda(x - x_{offset})]$, where we find $y_{min} \approx -0.00540$, $x_{offset} \approx 143$ years, and $\lambda \approx 0.0773\ \text{years}^{-1}$.

There is a paucity of public Australian data to inform the less-severe outcomes of ED and GP. National Centre for Immunisation Research and Surveillance (NCIRS) analysis estimated that roughly one in six Acute Respiratory Illness (ARI) ED presentations in adults aged over 65 in 2018/2019 led to hospital admission [70]. We therefore assumed that the probability of ED as a final endpoint is five-fold higher than the probability of at-least-hospitalisation. We benchmarked the probability of seeking care at a GP with reference to Korsten et al. 2021[71], a European cohort study, which found that the incidence of symptomatic RSV in adults aged over 75 is 4.2–7.2% per year. The above study also estimated the probability of presenting to GP given Acute Respiratory Tract Infection (ARTI) to be ~30%. Combining these pieces of evidence, we estimated that the incidence of GP visits per 100,000 in each age group over 60 per year should be around 2,000–3,000. That is, the probability of an individual seeking care at a GP *given infection* (the quantity of interest for the clinical pathways model) depended on the population-level incidence of infection per year. We therefore adjusted the probability of seeking medical care for adults aged ≥60 such that we recovered the appropriate number of GP visits per age group per year. We found that a sigmoid with functional form $y = \frac{y_{max} + y_{min}}{2} + \frac{y_{max} - y_{min}}{2} \tanh[a\ (age\ +\ b)]$, with parameters

0.5, 0.025, 0.05, −90 for $y_{max}$, $y_{min}$, $a$, and $b$, respectively, adequately recovers the desired GP incidence. We show all conditional transition probabilities in Figure 2, with associated data by which they are informed, where possible.

**Figure 2: Conditional transition probabilities for the clinical pathways model, and associated data with which they are informed. Abbreviations in legend: SMC = seek medical care, H⁺ = at-least hospitalisation, ICU⁺ = at-least ICU, F←ICU = death from ICU, F←H = death from hospitalisation (without ICU).**

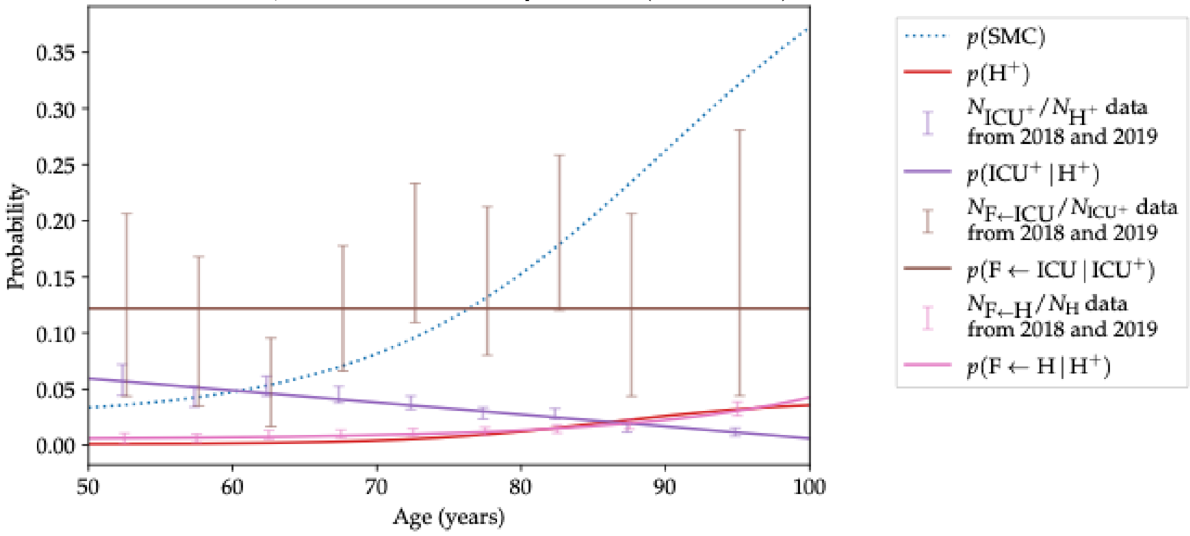


The method to convert the conditional to absolute probabilities of each endpoint (given infection) is described in the Supplementary Materials of Carlin et al 2025[26]. We show the resultant probabilities in Figure 3, with three panels that zoom in on different endpoints. Note that not-at-risk and at-risk individuals have differentiated absolute probabilities of each endpoint. The mathematical details are again described in Carlin et al 2025[26].

**Figure 3: Probability of different final clinical endpoints, given infection, as a function of age.** Panels on the right show zoomed regions of the main panel, to emphasise that all these probabilities are age- and risk-specific. The probabilities for at-risk individuals are shown as dashed curves, while not-at-risk individuals are shown as dotted curves, solid lines are for the weighted average of at-risk and not-at-risk individuals. Abbreviations: nSMC = does not seek medical care, GP = general practitioner visit, ED = emergency department, H = hospitalisation, ICU = intensive care unit admission, F←ICU = death from ICU, F←H = death from hospitalisation.

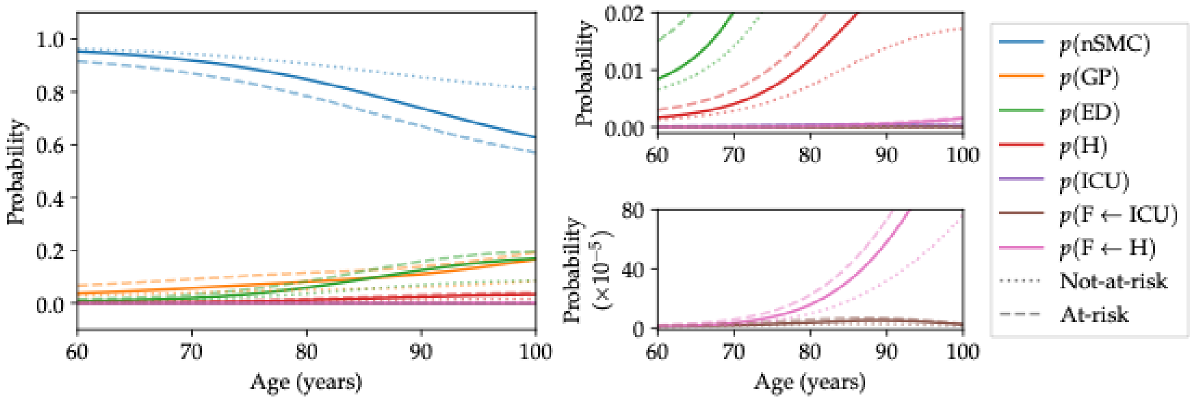


We tabulate in Table 2 the prevalence of the “at-risk” state (i.e. at-least one risk condition) assumed in the model, as a function of age.

**Table 2: Prevalence of at least one risk condition, as a function of age in the general population model**

| Age (years) | Prevalence (%) |
|---|---|
| 60 | 21.9 |
| 61 | 23.0 |
| 62 | 24.1 |
| 63 | 25.2 |
| 64 | 26.4 |
| 65 | 27.6 |
| 66 | 28.9 |
| 67 | 30.2 |
| 68 | 31.5 |
| 69 | 32.8 |
| 70 | 34.2 |
| 71 | 35.6 |
| 72 | 37.0 |
| 73 | 38.5 |
| 74 | 39.9 |
| 75 | 41.4 |

As described in the main text, all individuals aged ≥60 years in the Indigenous population model are considered "at-risk" for the purpose of vaccine eligibility, and have clinical endpoint multiplied by the numbers in Table 3 compared to an individual in the general population model, such that we find *post-facto* an IRR of 2.9 for each endpoint.

**Table 3: Necessary multipliers for Indigenous population model endpoint probabilities such that we find an IRR of 2.9 at each endpoint**. GP = general practitioner visit, ED = emergency department, H = hospitalisation, ICU = intensive care unit admission.

| Endpoint | Multiplier |
|---|---|
| GP | 3.53 |
| ED/H | 5.19 |
| ICU | 3.92 |
| Death | 6.31 |
