## Supplemental appendix 2 for "Cost-effectiveness of respiratory syncytial virus vaccination for older adults: a modelling analysis"

### Additional file 2: Further results

**Journal name:** Pharmacoeconomics.

Victoria L. Oliver[1]*, Julian B. Carlin[2]*, Yingying Wang[1]*, Violeta Spirkoska[2], Adrian Marcato[2], Kylie S. Carville[2,3], Robert Moss[1], David J. Price[1,2], Patricia T. Campbell[2], Jodie McVernon[2]^, Natalie Carvalho[1]^

*Joint first authors
^ Joint senior authors

[1] Melbourne School of Population and Global Health, University of Melbourne, VIC, Australia
[2] Department of Infectious Diseases, The University of Melbourne at the Peter Doherty Institute for Infection and Immunity, Victoria, Australia
[3] Victorian Infectious Diseases Reference Laboratory, The Royal Melbourne Hospital at the Peter Doherty Institute for Infection and Immunity, Victoria, Australia

**Figure 1: Flow chart of studies identified, screened, and included from the literature review.**

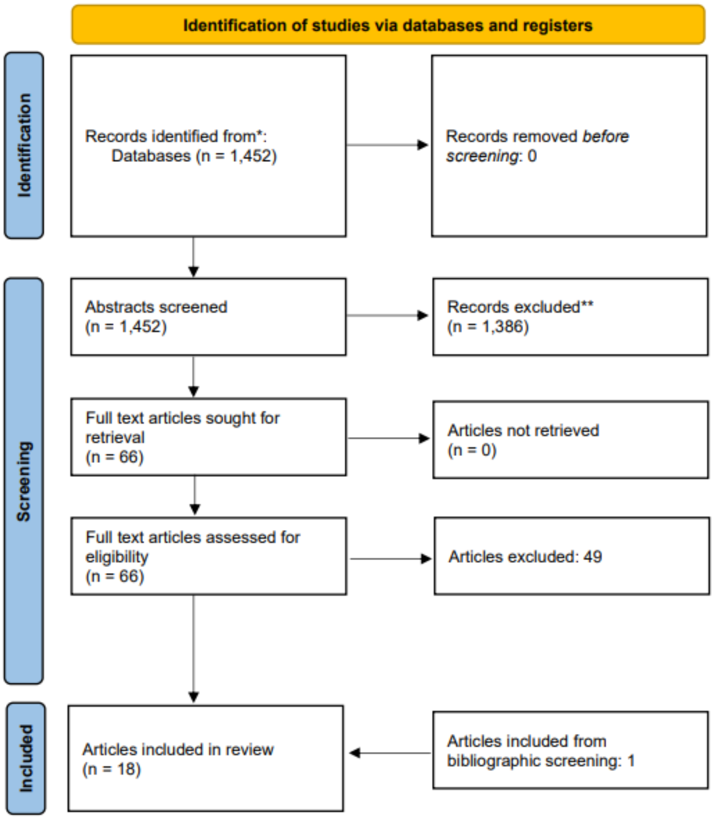

**Table 1: Summary of key model inputs and findings of published cost-effectiveness analyses. Our study has been included here to facilitate comparison.**

| Study, analysis | Country, perspective | Baseline incidence of hospitalisation (per 100,000)[a] | Hospitalised case-fatality risk [a] | Mortality rate (per 100,000)[a] | Duration of protection (against hospitalisation) | Vaccine price (2024 USD)[b] | Number needed to vaccinate to avoid 1 hospitalisation or death | Cost-effectiveness of vaccination strategy[c], ICER (2024 USD/QALY)[b] | WTP threshold (2024 USD/QALY)[b] |
|---|---|---|---|---|---|---|---|---|---|
| **Analyses of the cost effectiveness of vaccination for adults aged ≥60 or ≥65 years compared to no vaccination** | | | | | | | | | |
| **This study** | Australia, Healthcare | 491.3 | 1.7% | 8.4 | Waning to reach 0% at 36 months | 63 | Hospitalisation: 483<br>Death: 26,220 | Not cost-effective, 192,202 | 10,000 - 33,000 |
| **Meijboom et. al., 2013 [59]** | Netherlands, Healthcare[d] | 46.2[e] | 46.0% (≥65) | 21.3 (≥65) | 1 season (no waning) | 66 | NR | Not cost-effective, 67,878 | 66,000 |
| **Ortega-Sanchez, 2023 [24], UM-CDC model - Abrysvo** | USA, Societal | 162 (≥65) | Not reported | Unknown | Waning to 0% at 24 months | 200 | NR | Not cost-effective, 118,735 | 102,000 |
| **Ortega-Sanchez, 2023 [24], UM-CDC model - Arexvy** | USA, Societal | 162 (≥65) | Not reported | Unknown | Waning to 0% at 24 months | 270 | NR | Not cost-effective, 205,638 | 102,000 |
| **Hutton et. al., 2024 [62], Abrysvo** | USA, Societal | 65.5 (60–64)<br>93.8 (65–69)<br>118.7 (70–74)<br>302.9 (≥75) | 3.9% (60–64)<br>4.3% (65–74)<br>5.7% (≥75) | 2.6 (60–64)<br>4.0 (65–69)<br>5.1 (70–74)<br>17.3 (≥75) | Waning over 14 months | 295 | Hospitalisation: 440<br>Death: 9,385 | Not cost-effective, 196,842 | 125,000–190,000 |
| **Hutton et. al., 2024 [62], Arexvy** | USA, Societal | 65.5 (60–64)<br>93.8 (65–69)<br>118.7 (70–74)<br>302.9 (≥75) | 3.9% (60–64)<br>4.3% (65–74)<br>5.7% (≥75) | 2.6 (60–64)<br>4.0 (65–69)<br>5.1 (70–74)<br>17.3 (≥75) | Waning over 18 months | 280 | Hospitalisation: 478<br>Death: 8,656 | Cost-effective, 176,557 | 125,000–190,000 |
| **Komiya et. al., 2025 [22]** | Japan, Healthcare & Societal | 413.1 | 13.6% | 56.2 | Waning to 0% over 42 months | 158 | NR | Cost-effective, 9,693 (healthcare) 5,967 (societal) | 33,000 |
| **Averin et. al. 2025 [57]** | Germany, Healthcare & Societal | 144.2 (60–69)[e]<br>433.5 (70–79)[e]<br>876.4 (≥80)[e] | 7.8% | 11.2 (60–69)<br>33.8 (70–79)<br>68.4 (≥80) | Waning to 0% over 42 months | 230 | NR | Cost-effective, 42,311 (healthcare) 39,035 (societal) | 13,00–54,00 |
| **Gourzoulidis et. al., 2024 [44]** | Greece, Public payer | 206.6 (60–64)[e]<br>408.3 (65–74)[e]<br>645.5 (75–84)[e]<br>921.9 (≥85)[e] | 7.4% | 15.3 (60–64)<br>30.2 (65–74)<br>47.8 (75–84)<br>68.2 (≥85) | Waning over 41 months | 223 | NR | Cost-effective, 21,438 | 46,625 |

| Study, analysis | Country, perspective | Baseline incidence of hospitalisation (per 100,000)[a] | Hospitalised case-fatality risk [a] | Mortality rate (per 100,000)[a] | Duration of protection (against hospitalisation) | Vaccine price (2024 USD)[b] | Number needed to vaccinate to avoid 1 hospitalisation or death | Cost-effectiveness of vaccination strategy[c], ICER (2024 USD/QALY)[b] | WTP threshold (2024 USD/QALY)[b] |
|---|---|---|---|---|---|---|---|---|---|
| **La et. al., 2024 [60]** | USA, Societal | 338.7 | 5.15% | 17.4 | Waning to 28.2% at 60 months | 294 | Hospitalisation: 182 Death: 2,279 | Cost-effective, 18,430 | 50,000 |
| **Mizukami et. al., 2024 [23]** | Japan, Healthcare & Societal | 407.6 | 4.77% | 19.4 | Waning over 2 seasons | 132 | NR | Cost-effective, 27,616 (healthcare) 26,703 (societal) | 33,000 – 39,000 |
| **Ortega-Sanchez 2023 [24], Pfizer model** | USA, Societal | 300 (≥65) | Not reported | Unknown | Waning over 43 months | 270 | NR | Cost-effective, 23,921 | 102,000 |
| **Ortega-Sanchez 2023 [24], GSK model** | USA, Societal | 256.3 (≥65) | Not reported | Unknown | Waning over 24 months | 200 | NR | Cost-effective, 64,348 | 102,000 |
| **Gessner et. al., 1999 [66]** | USA, Societal | 44.0 (≥65) | 11% (≥65) | 4.8 (≥65) | 1 season (no waning) | 52 | NR | Cost-effective, 5,342 | 35,000 |
| **Wang et. al., 2023 [46], Abrysvo** | Hong Kong, Public healthcare provider | 141.45 | 7.7% | 10.9 | Waning to 75% in season two | 50[f] | NR | Cost-effective, 26,843 | 51,000 |
| **Wang et. al., 2023 [46], Arexvy** | Hong Kong, Public healthcare provider | 141.45 | 7.7% | 10.9 | Waning to 52.9% in season two | 67.5[f] | NR | Cost-effective, 48,633 | 51,000 |
| **Zeevat et. al., 2022 [58]** | Netherlands, Societal | 51.1 | 38.6% | 19.7 | 1 year (no waning) | NMB at dose price of 17–53 USD[h] | NR | Dose price dependent | 22,000–55,000 |

| Study, analysis | Country, perspective | Baseline incidence of hospitalisation (per 100,000)[a] | Hospitalised case-fatality risk [a] | Mortality rate (per 100,000)[a] | Duration of protection (against hospitalisation) | Vaccine price (2024 USD)[b] | Number needed to vaccinate to avoid 1 hospitalisation or death | Cost-effectiveness of vaccination strategy[c], ICER (2024 USD/QALY)[b] | WTP threshold (2024 USD/QALY)[b] |
|---|---|---|---|---|---|---|---|---|---|
| **Hodgson et al. 2020 [28]** | USA, Public healthcare provider | 70 (65–74)<br>245 (≥75) | 10% (65–74)<br>15% (≥75) | 7.0 (65–74)<br>36.8 (≥75) | 1 year (no waning) | NMB at dose price of 27 USD | NR | Dose price dependent | 34,000 |
| **Shoukat et. al., 2024 [35], Abrysvo** | Canada, Healthcare & Societal | 145 | 7.6% (60–69)<br>8.1% (70–79)<br>14% (≥80) | 11.0 (60–69)<br>11.7 (70–79)<br>20.3 (≥80) | Waning over 24 months | NMB at dose price of 107-134 USD[g] | NR | Dose price dependent | 36,000 |
| **Shoukat et. al., 2024 [35], Arexvy** | Canada, Healthcare & Societal | 145 | 7.6% (60–69)<br>8.1% (70–79)<br>14% (≥80) | 11.0 (60–69)<br>11.7 (70–79)<br>20.3 (≥80) | Waning over 24 months | NMB at dose price of 104-137 USD[g] | NR | Dose price dependent | 36,000 |
| **Moghadas et. al., 2023 [63], Abrysvo** | USA, Societal | 214 | 8.8%[i] | 18.8 | Waning over 24 months | NMB at dose price of 197 – 245 USD[j] | Hospitalisation: 281-326[j]<br>Death: 3,384-3,977[j] | Dose price dependent | 97,000 |
| **Moghadas et. al., 2023 [63], Arexvy** | USA, Societal | 214 | 8.8%[i] | 18.8 | Waning over 24 months | NMB at dose price of 210 – 235 USD[j] | Hospitalisation: 301-325<br>Death: 3,536-3,984 | Dose price dependent | 97,000 |
| **Zeevat et. al., 2022 [58]** | UK, Third party payer | 151.4 (≥65) | 59.7% (≥65) | 90.4 (≥65) | 1 year (no waning) | NMB at dose price of 89–135 USD[h] | NR | Dose price dependent | 25,000–39,000 |
| **Herring et. al., 2022 [65]** | USA, Third party payer | 335[k] | 8.7% | 29.1 | 1 season (no waning) | NMB at dose price of 152 – 299 USD[h] | Hospitalisation: 569<br>Death: 3,113 | Dose price dependent | 53,000 – 106,000 |
| **Analyses of the cost effectiveness of vaccination for adults aged ≥60 or ≥65 years compared to next best RSV vaccination strategy for adults** | | | | | | | | | |
| **Rudd et. al., 2025 [61] – Public Health Agency of Canada model - Abrysvo** | Canada, Health sector | 165.4[e] | 6.6% | 10.9 | Waning over 36 months | 168 | NR | Dominated (next best strategy not reported) | 37,000 |

| Study, analysis | Country, perspective | Baseline incidence of hospitalisation (per 100,000)[a] | Hospitalised case-fatality risk [a] | Mortality rate (per 100,000)[a] | Duration of protection (against hospitalisation) | Vaccine price (2024 USD)[b] | Number needed to vaccinate to avoid 1 hospitalisation or death | Cost-effectiveness of vaccination strategy[c], ICER (2024 USD/QALY)[b] | WTP threshold (2024 USD/QALY)[b] |
|---|---|---|---|---|---|---|---|---|---|
| **Rudd et. al., 2025 [61] – Public Health Agency of Canada model - Arexvy** | Canada, Health sector | 165.4[e] | 6.6% | 10.9 | Waning over 36 months | 168 | NR | Dominated (next best strategy not reported) | 37,000 |
| **Rudd et. al., 2025 [61] – Pfizer model - Abrysvo** | Canada, Health sector | 165.4[e] | 6.6% | 10.9 | Waning over 36 months | 168 | NR | Dominated (next best strategy not reported) | 37,000 |
| **Rudd et. al., 2025 [61] – Pfizer model - Arexvy** | Canada, Health sector | 165.4[e] | 6.6% | 10.9 | Waning over 36 months | 168 | NR | Dominated (next best strategy not reported) | 37,000 |
| **Rudd et. al., 2025 [61] – GSK model - Abrysvo** | Canada, Health sector | 165.4[e] | 6.6% | 10.9 | Waning over 36 months | 168 | NR | Dominated (next best strategy not reported) | 37,000 |
| **Rudd et. al., 2025 [61] – GSK model - Arexvy** | Canada, Health sector | 165.4[e] | 6.6% | 10.9 | Waning over 36 months | 168 | NR | Dominated (next best strategy not reported) | 37,000 |
| **Li et. al., 2025, Denmark** | Denmark, Healthcare & Societal | 63.3 - 173.7 [e,l] | 7.13 | 4.5 – 12.4 | Waning over 24 months | 162 | NR | Dependent on WTP | Cost-effective compared to vaccination of ≥75-year-olds at WTP = 70,000 [m] |
| **Li et. al., 2025, Finland** | Finland, Healthcare & Societal | 493.3 - 637.5 [e,l] | 7.13 | 35.2 – 45.5 | Waning over 24 months | 162 | NR | Not cost-effective compared to vaccination of ≥75 year old | 162,000 |
| **Li et. al., 2025, Netherlands** | Netherlands, Healthcare & Societal | 213.1 - 859.7 [e,l] | 7.13 | 15.2 – 61.3 | Waning over 24 months | 162 | NR | Dependent on WTP | Cost-effective compared to vaccination of ≥75-year-olds |

| Study, analysis | Country, perspective | Baseline incidence of hospitalisation (per 100,000)[a] | Hospitalised case-fatality risk [a] | Mortality rate (per 100,000)[a] | Duration of protection (against hospitalisation) | Vaccine price (2024 USD)[b] | Number needed to vaccinate to avoid 1 hospitalisation or death | Cost-effectiveness of vaccination strategy[c], ICER (2024 USD/QALY)[b] | WTP threshold (2024 USD/QALY)[b] |
|---|---|---|---|---|---|---|---|---|---|
| | | | | | | | | | at WTP = 76,000[m] |
| **Li et. al., 2025, Spain** | Valencia - Spain, Healthcare & Societal | 427.7 – 836.4 [e,l] | 7.13 | 30.5 – 59.6 | Waning over 24 months | 162 | NR | Dependent on WTP | Cost-effective compared to vaccination of ≥75-year-olds at WTP = 126,000 [m] |
| **Tuite et. al., 2024- Abrysvo** | Canada, Healthcare & Societal | 71.25 (60-69)[e]<br>144.6 (70-79)[e]<br>461.1 (80+)[e] | 6.6% (65-74)<br>10.1% (75+) | Unknown | Waning over 24 months | 168 | Hospitalization: 338, Death: 4,315 | Dominated[n] | NR |
| **Tuite et. al., 2024- Arexvy** | Canada, Healthcare & Societal | 71.25 (60-69)[e]<br>144.6 (70-79)[e]<br>461.1 (80+)[e] | 6.6% (65-74)<br>10.1% (75+) | Unknown | Waning over 24 months | 168 | Hospitalization: 353, Death: 3,973 | Dominated[n] | NR |
| **Analyses of the cost effectiveness of vaccination for adults with elevated risk aged ≥60 or 65 years compared to no vaccination** | | | | | | | | | |
| **This study** | Australia, Healthcare | 1423.0 | 1.7% | 24.2 | 36 months | 63 | Hospitalisation: 104<br>Death: 5,815 | Cost-saving | 10,000 - 33,000 |
| **Averin et. al. 2025 [57]** | Germany, Healthcare & Societal | 202.6 (60–69)[e]<br>501.5 (70–79)[e]<br>930.3 (≥80)[e] | 6.1% (60–69)<br>9.0% (70–79)<br>10.6% (≥80) | 12.4 (60–69)<br>45.1 (70–79)<br>98.6 (≥80) | Waning to 0% over 42 months | 230 | NR | Cost-effective, 34,998 (healthcare)<br>31,728 (societal) | 13,000–54,000 |
| **Gourzoulidis et. al., 2024 [44]** | Greece, Public payer | 373 (60–64)[e]<br>558 (65–74)[e]<br>740 (75–784)[e]<br>977 (≥85)[e] | 7.4% | 27.6 (60–64)<br>41.3 (65–74)<br>54.8 (75–84)<br>72.3 (≥85) | Waning over 41 months | 223 | NR | Cost-effective, 15,242 | 47,000 |
| **Meijboom et. al., 2013 [59]** | Netherlands, Healthcare[d] | 88.2 (60–64)[e]<br>93.0 (65–74)[e]<br>188.3 (75–84)[e]<br>395.5 (≥85)[e] | 23.7% (60–64)<br>22.5% (65–74)<br>34.4% (75–84)<br>53.3% (≥85) | 20.9 (60–74)<br>64.7 (75–84)<br>210.9 (≥85) | 1 season (no waning) | 66 | NR | Cost-effective, 31,805 | 87,000 |
| **Shoukat et. al., 2024 [35], Abrysvo** | Canada, Healthcare & Societal | 330.6[e] | 7.6% (60–69)<br>8.1% (70–79)<br>14% (≥80) | 25.1 (60–69)<br>26.8 (70–79)<br>46.3 (≥80) | Waning over 24 months | NMB at dose price | NR | Price dependent | 36,000 |

| Study, analysis | Country, perspective | Baseline incidence of hospitalisation (per 100,000)[a] | Hospitalised case-fatality risk [a] | Mortality rate (per 100,000)[a] | Duration of protection (against hospitalisation) | Vaccine price (2024 USD)[b] | Number needed to vaccinate to avoid 1 hospitalisation or death | Cost-effectiveness of vaccination strategy[c], ICER (2024 USD/QALY)[b] | WTP threshold (2024 USD/QALY)[b] |
|---|---|---|---|---|---|---|---|---|---|
| | | | | | | of 127 USD[g] | | | |
| **Shoukat et. al., 2024 [35], Arexvy** | Canada, Healthcare & Societal | 330.6[e] | 7.6% (60–69)<br>8.1% (70–79)<br>14% (≥80) | 25.1 (60–69)<br>26.8 (70–79)<br>46.3 (≥80) | Waning over 24 months | NMB at dose price of 119 USD[g] | NR | Price dependent | 36,000 |
| **Analyses of the cost effectiveness of vaccination for adults with elevated risk aged ≥60 or 65 years compared to next best RSV vaccination strategy** | | | | | | | | | |
| **Rudd et. al., 2025 [61] – Public Health Agency of Canada model - Abrysvo** | Canada, Health sector | 165.4[e] | 6.6% | 10.9 | Waning over 36 months | 168 | NR | Not cost-effective or extendedly dominated[o] | 37,000 |
| **Rudd et. al., 2025 [61] – Public Health Agency of Canada model - Arexvy** | Canada, Health sector | 165.4[e] | 6.6% | 10.9 | Waning over 36 months | 168 | NR | Not cost-effective or extendedly dominated[o] | 37,000 |
| **Rudd et. al., 2025 [61] – Pfizer model - Abrysvo** | Canada, Health sector | 165.4[e] | 6.6% | 10.9 | Waning over 36 months | 168 | NR | Not cost-effective[p] | 37,000 |
| **Rudd et. al., 2025 [61] – Pfizer model - Arexvy** | Canada, Health sector | 165.4[e] | 6.6% | 10.9 | Waning over 36 months | 168 | NR | Not cost-effective[p] | 37,000 |
| **Rudd et. al., 2025 [61] – GSK model - Abrysvo** | Canada, Health sector | 165.4[e] | 6.6% | 10.9 | Waning over 36 months | 168 | NR | Not cost-effective or extendedly dominated[o] | 37,000 |
| **Rudd et. al., 2025 [61] – GSK model - Arexvy** | Canada, Health sector | 165.4[e] | 6.6% | 10.9 | Waning over 36 months | 168 | NR | Not cost-effective or extendedly dominated[o] | 37,000 |

| Study, analysis | Country, perspective | Baseline incidence of hospitalisation (per 100,000)[a] | Hospitalised case-fatality risk [a] | Mortality rate (per 100,000)[a] | Duration of protection (against hospitalisation) | Vaccine price (2024 USD)[b] | Number needed to vaccinate to avoid 1 hospitalisation or death | Cost-effectiveness of vaccination strategy[c], ICER (2024 USD/QALY)[b] | WTP threshold (2024 USD/QALY)[b] |
|---|---|---|---|---|---|---|---|---|---|
| **Tuite et. al., 2024- Abrysvo** | Canada, Healthcare & Societal | 71.25 (60-69)[e]<br>144.6 (70-79)[e]<br>461.1 (80+)[e] | 6.6% (65-74)<br>10.1% (75+) | Unknown | Waning over 24 months | 168 | Hospitalization: 338, Death: 4,315 | Not cost-effective or extendedly dominated[o] | NR |
| **Tuite et. al., 2024- Arexvy** | Canada, Healthcare & Societal | 71.25 (60-69)[e]<br>144.6 (70-79)[e]<br>461.1 (80+)[e] | 6.6% (65-74)<br>10.1% (75+) | Unknown | Waning over 24 months | 168 | Hospitalization: 353, Death: 3,973 | Not cost-effective or extendedly dominated[o] | NR |

NMB, net monetary benefit; NR, not reported; USD, United States dollars; WTP, Willingness to pay;

[a] Unless otherwise stated, hospitalisation incidence, hospitalised case fatality rate, and mortality rate have been reported for adults ≥60 years in the population eligible for vaccination (general population or adults with elevated risk). Mortality rate has been calculated based on hospitalisation incidence and hospitalised case fatality rate.

[b] Dose price, ICER and WTP thresholds reported in local currency units have been converted to USD using exchange rates for the year of the analysis. Dose price, ICER and WTP have been adjusted to 2024 USD using GDP deflators to express all values in 2024 USD. Thresholds are expressed to the nearest 1,000 USD

[c] Where results from multiple scenario analyses were presented in the published study, ICER for the scenario most closely aligned with our study has been presented. Assessment of cost-effectiveness presented in the table was made based on upper WTP threshold.

[d] Perspective adopted was not explicitly reported, healthcare sector perspective assumed based on included costs

[e] Hospitalisation incidence has been computed based on data reported for those with and without risk conditions in the article.

[f] Four price estimates were explored for each vaccine. Results are reported for the estimate most comparable to the price used in our analysis.

[g] Upper and lower NMB bounds represent results for healthcare and societal perspective respectively

[h] NMB ranges correspond to lower and upper WTP threshold used

[i] Computed from uniform range

[j] Ranges encompass results for different waning curves. Results reflect the two-year horizon scenario analysis.

[k] Study presented alternative analysis scenarios based on two different data sources for hospitalisation incidence. Analysis with hospitalisation incidence most comparable to our study has been presented.

[l] Range represent inputs from three different methods for estimating hospitalisation incidence.

[m] Vaccination of adults ≥65 years was preferred over vaccination of adults ≥75 years for only one estimate of hospitalisation incidence modelled but was not preferred up to a WTP threshold of 162,000 USD otherwise

[n] Vaccination of adults ≥60 years was dominated by vaccination of adults ≥70 years along with at-risk adults ≥50 years. Vaccination of adults ≥65 years was dominated by vaccination of adults ≥75 years along with at-risk adults ≥60 years

[o] Vaccination of at-risk adults ≥60 years was not cost-effective compared to vaccination of at-risk adults ≥70 years. Vaccination of at-risk adults ≥65 years was extendedly dominated.

[p] Vaccination of at-risk adults ≥60 years was not cost-effective compared to vaccination of at-risk adults ≥65 years. Vaccination of at-risk adults ≥65 years was not cost-effective compared to vaccination of at-risk adults ≥70 years.

**Table 2: Clinical outcome results for all strategies simulated. All numbers are scaled to a population of 100,000 adults ≥60 years of age. Data show the average number of clinical outcomes in the no vaccination strategy and the number of outcomes averted by each vaccination strategy.**

| Vaccination strategy | Doses delivered | GP | ED | H | ICU | Deaths |
|---|---|---|---|---|---|---|
| **General population** | | | | | | |
| No intervention, number of cases | | 4530.8 (4263–4847) | 2347.6 (2162–2543) | 470.3 (391–551) | 12.5 (2–27) | 8.6 (1–21) |
| Year-round vaccination of at-risk ≥60 with others ≥75, number of cases averted | 35,343 | 681.3 (532–834) | 483.8 (391–584) | 96.6 (61–137) | 2.4 (–6–11) | 1.9 (0–6) |
| Seasonal vaccination of at-risk ≥60, others ≥75, number of cases averted | 34,533 | 763.3 (598–937) | 544.0 (438–659) | 108.9 (71–151) | 2.6 (–6–12) | 2.2 (0–6) |
| Seasonal vaccination of at-risk ≥60 with others ≥70, number of cases averted | 42,527 | 859.0 (683–1050) | 593.1 (480–716) | 118.3 (80–165) | 2.9 (–6–13) | 2.3 (0–7) |
| Seasonal vaccination of at-risk ≥60 with others ≥65, number of cases averted | 52,774 | 973.6 (791–1173) | 631.1 (513–754) | 126.1 (85–170) | 3.2 (–6–14) | 2.4 (0–10) |
| Seasonal vaccination of ≥60, number of cases averted | 65,549 | 1128.5 (933–1347) | 676.6 (556–810) | 135.6 (94–184) | 3.5 (–6–14) | 2.5 (0–10) |
| Seasonal vaccination of ≥75, number of cases averted | 21,032 | 432.2 (306–561) | 415.6 (325–509) | 83.1 (52–118) | 1.7 (–18–43) | 1.8 (0–5) |
| **Indigenous population** | | | | | | |
| No intervention, number of cases | | 13145.4 (12405–13916) | 6814.2 (6302–7329) | 1363.4 (1151–1585) | 35.3 (5–74) | 24.3 (2–56) |
| Seasonal vaccination of ≥60, number of cases averted | 37,795 | 3000.0 (2451–3603) | 1803.9 (1473–2167) | 362.0 (248–483) | 9.2 (–15–38) | 6.5 (1–26) |

GP = general practitioner visit, ED = emergency department, H = hospitalisation, ICU = intensive care unit admission.

**Table 3: Number needed to vaccinate to avert each clinical outcome compared to the no-intervention strategy.**

| Vaccination strategy | GP | ED | H | ICU | Death |
|---|---|---|---|---|---|
| **General population** | | | | | |
| Year-round vaccination of at-risk ≥60 with others ≥75 | 52 | 73 | 366 | 14,726 | 18,602 |
| Seasonal vaccination of at-risk ≥60, others ≥75 | 45 | 63 | 317 | 13,282 | 15,697 |
| Seasonal vaccination of at-risk ≥60 with others ≥70 | 50 | 72 | 359 | 14,664 | 18,490 |
| Seasonal vaccination of at-risk ≥60 with others ≥65 | 54 | 84 | 419 | 16,492 | 21,989 |
| Seasonal vaccination of ≥60 | 58 | 97 | 483 | 18,728 | 26,220 |
| Seasonal vaccination of ≥75 | 49 | 51 | 253 | 12,372 | 11,684 |
| **Indigenous population** | | | | | |
| Seasonal vaccination of ≥60 | 13 | 21 | 104 | 4108 | 5,815 |

GP = general practitioner visit, ED = emergency department, H = hospitalisation, ICU = intensive care unit admission.

**Figure 2: Cost-effectiveness planes from probabilistic sensitivity analysis of vaccination strategies for the general population (top) and the Indigenous population (bottom) compared to no vaccination.** For the general population, vaccination of adults aged ≥75 years and at-risk adults aged ≥60 years was modelled. For the Indigenous population, vaccination of adults aged ≥60 years was modelled.

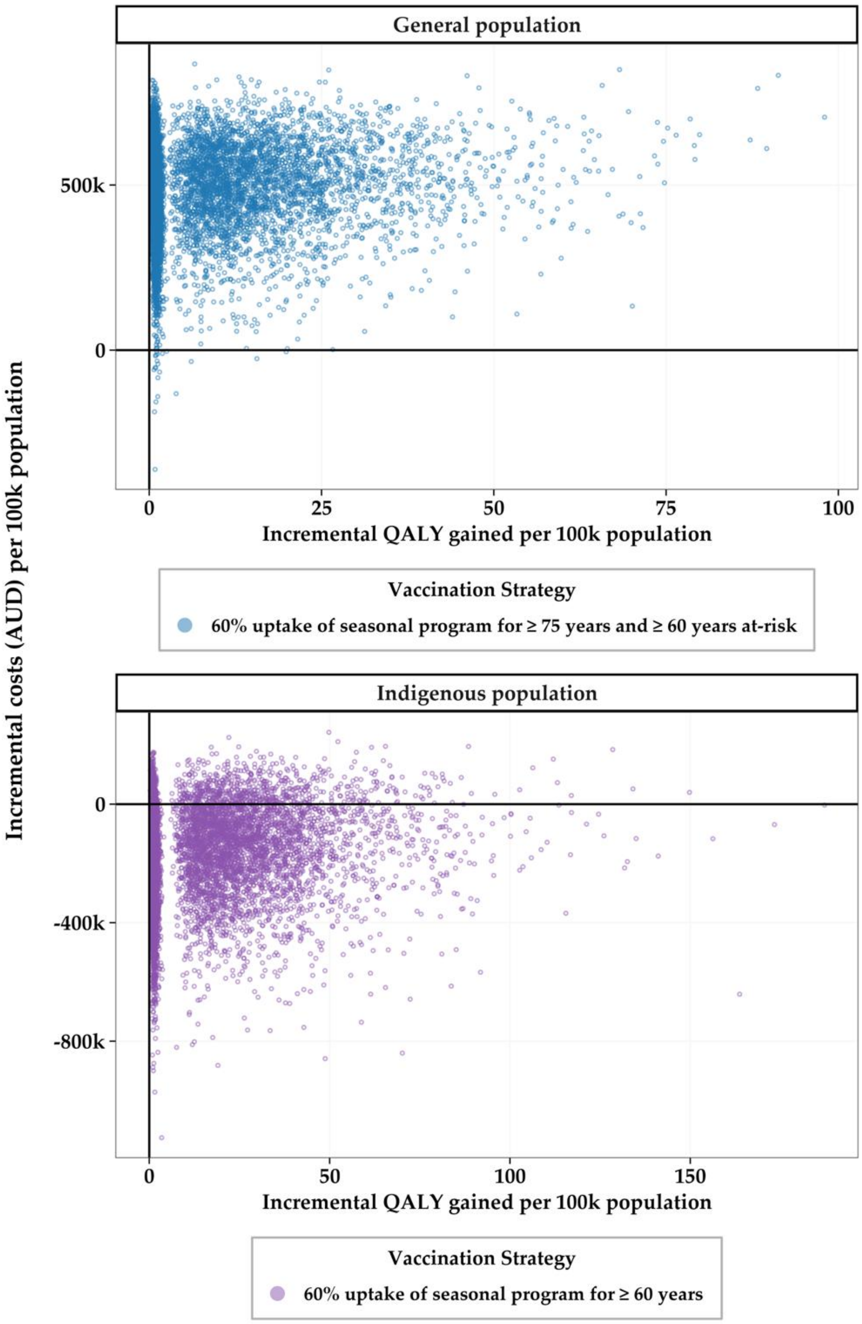
